# Multi-ancestry Multivariate Genome-Wide Analysis Highlights the Role of Common Genetic Variation in Cardiac Structure, Function, and Heart Failure-related Traits

**DOI:** 10.1101/2021.08.03.21261508

**Authors:** Michael G. Levin, Noah L. Tsao, Tiffany R. Bellomo, William P. Bone, Krishna G. Aragam, Yifan Yang, Michael P. Morley, Megan Burke, Renae L. Judy, Zoltan Arany, Thomas P. Cappola, Sharlene M. Day, Patrick T. Ellinor, Kenneth B. Margulies, Benjamin F. Voight, Scott M. Damrauer

## Abstract

Heart failure (HF) is a leading cause of cardiovascular morbidity and mortality, yet the contribution of common genetic variation to HF risk has not been fully elucidated, particularly in comparison to other common cardiometabolic traits. We conducted a multi-ancestry genome-wide association study (GWAS) meta-analysis of all-cause HF including up to 56,722 HF cases and 1,133,054 controls, identifying 4 novel loci. We then performed a multi-ancestry multivariate association study of HF and related cardiac imaging endophenotypes, identifying 71 conditionally-independent variants, including 16 novel loci. Secondary colocalization and transcriptome-wide association analyses identified known and novel candidate cardiomyopathy genes, which were validated in gene-expression profiling of failing and healthy human hearts. Colocalization, gene expression profiling, and Mendelian randomization provided convergent evidence for the roles of *BCKDHA* and circulating branch-chain amino acids in heart failure and cardiac structure. Finally, proteome-wide Mendelian randomization revealed 11 circulating proteins associated with HF or quantitative imaging traits. These analyses highlight similarities and differences among heart failure and associated cardiovascular imaging endophenotypes, implicate novel common genetic variation in the pathogenesis of HF, and identify circulating proteins that may represent novel cardiomyopathy treatment targets.

## INTRODUCTION

Heart failure (HF) is a common cardiovascular syndrome characterized by symptoms including shortness of breath, volume-overload, and functional limitation that result from structural or functional impairment of ventricular filling or ejection of blood^1–4^. HF affects > 38 million individuals globally, with rapidly growing prevalence, and is a primary cause of cardiovascular morbidity, mortality, hospitalization, and healthcare costs^5^. Despite the high population prevalence of HF, the role of common genetic variation in HF risk remains poorly understood. In comparison to other common cardiometabolic traits like coronary artery disease (CAD), myocardial infarction, diabetes, blood pressure, and obesity, where hundreds of genetic loci have been associated with disease risk, discovery of common genetic sequence variants associated with HF has been modest, with only 11 genomic loci identified in the largest genome-wide association study (GWAS) of HF to date^6^.

Several strategies have been described to improve power for GWAS locus discovery. Simulation and empiric studies have identified power gains of multiethnic GWAS^7^. A recent trans-ancestry genome-wide analysis of CAD combining European and Japanese studies identified 40 novel loci, bringing the total number of CAD-associated loci to 170^8^. In the context of related or correlated traits, multivariate scans have been employed, with a recent multivariate genome-wide-association meta-analysis (GWAMA) demonstrating a 26% increase in the number of variants associated with traits related to mood and affect^9^. HF is a heterogeneous clinical syndrome^10^, and imprecision in clinical phenotyping may represent a limitation to detecting common variant associations^11^. A recent multi-trait analysis jointly considered hypertrophic cardiomyopathy, dilated cardiomyopathy, and cardiac imaging traits, successfully identifying novel common genetic variants associated with these traits^12^. We hypothesized that jointly considering HF and related continuous quantitative cardiac imaging phenotypes may help resolve phenotypic imprecision and similarly improve power for genetic discovery.

In this study, we performed a multi-ancestry meta-analysis of HF genome-wide association studies to estimate the effect of common genetic sequence variants on all-cause HF risk. We then integrated multi-ancestry GWAS of cardiac imaging phenotypes and HF within the multivariate GWAMA framework to further improve power for novel locus discovery. Finally, we evaluated the genetic evidence for these associations using colocalization, transcriptome-wide association, gene-expression profiling, and Mendelian randomization. In summary, this study identifies novel HF risk variants and putative effector genes, prioritizes relevant tissues, highlights roles for common genetic sequence variation in the pathogenesis of HF and related traits, and identifies circulating proteins associated with HF and cardiovascular imaging phenotypes.

## RESULTS

### Multi-ancestry HF meta-analysis identifies 4 new loci

We conducted a multi-ancestry GWAS meta-analysis of all-cause HF to increase power for detection of HF-associated genetic variants. Combining two large-scale GWAS of all-cause HF from the HERMES consortium (European-ancestry) and BioBank Japan (BBJ; Japanese-ancestry), we analyzed 56,722 HF cases (HERMES N = 47,309; BBJ N = 9,413) and 1,133,054 controls (HERMES N = 930,014; BBJ N = 203,040). Fixed-effects inverse variance-weighted (IVW) meta-analysis was performed for 8,747,146 common (minor allele frequency [MAF] > 0.01) variants. We identified 15 loci where genetic associations reached the genome-wide significance (GWS) threshold (p < 5 × 10^−8^) (**SUPPLEMENTAL FIGURES 1-2**; **SUPPLEMENTAL TABLE 1**). Conditional analysis was performed at the 15 GWS loci, identifying 19 independent signals surrounding the lead variant at each locus (**SUPPLEMENTAL TABLE 2**). Four of the loci had not been previously reported in either the HERMES or BBJ HF GWAS (**TABLE 1**).

**TABLE 1:** Novel HF Loci.

| Chromosome | Position | SNP | Effect Allele | Other Allele | Effect | SE | P | Direction |
| --- | --- | --- | --- | --- | --- | --- | --- | --- |
| 4 | 45186139 | rs10938398 | a | g | 0.0407 | 0.0072 | 1.83E-08 | ++ |
| 6 | 22592216 | rs765782 | t | c | -0.045 | 0.0079 | 1.42E-08 | -- |
| 9 | 97619090 | rs10993372 | a | t | -0.04 | 0.0072 | 3.44E-08 | -- |
| 17 | 53400110 | rs12940636 | t | c | 0.0406 | 0.0074 | 4.40E-08 | ++ |

Following conditional analysis, 4 novel loci associated with HF were identified. These loci had not previously been identified in the HERMES or BBJ HF GWAS. SE = standard error, P = P-value.

We sought replication of independent significant variants in data from the release 4 FinnGen consortium HF GWAS (http://r4.finngen.fi/pheno/I9_HEARTFAIL_ALLCAUSE), which included 17,387 all-cause HF cases and 159,058 controls. Of the 19 conditionally-independent signals identified in the HERMES + BBJ meta-analysis, all lead variants (or proxies) were available for comparison in FinnGen (**SUPPLEMENTAL TABLE 3**). In total, 14 of 19 SNP-disease associations had the same direction of effect (73.7% concordance; binomial test with one-tailed p = 0.032). All 6 SNP-trait associations with p < 0.05 in FinnGen had concordant direction of effect (binomial test with one-tailed p = 0.016).

To determine associations between the novel HF loci and other traits, we queried summary-level results from 34,513 GWAS collected by the Open GWAS Project. The lead variants at the novel HF loci were associated with several obesity-related traits (Eg. body mass index, fat mass), diabetes, and atrial fibrillation among others (**SUPPLEMENTAL TABLE 4**), representing known and common HF risk factors.

### Heart failure and cardiac structure/function phenotypes are genetically correlated

Clinically, symptoms of heart failure are often linked to abnormalities of cardiac structure/function. Cross-trait linkage disequilibrium score regression (LDSC) was performed to estimate the genetic correlation of HF with previously-reported cardiac imaging-derived measures of cardiac structure and function^13,14^ including: cardiac MRI-derived measures of left-ventricular end-diastolic volume (LVEDV_MRI_), left-ventricular end-systolic volume (LVESV_MRI_), and left-ventricular ejection fraction (LVEF_MRI_) obtained from a GWAS of cardiac MRI traits among 36,041 healthy UK Biobank participants^15^, and transthoracic echocardiography (TTE)-derived measures of left ventricular internal diameter end-diastole (LVIDd_TTE_), left ventricular internal diameter end-systole (LVIDs_TTE_), and LVEF_TTE_ obtained from a GWAS among up to 19,676 participants^16^. Although the cardiac MRI GWAS were performed in healthy participants, HF was significantly correlated with all imaging phenotypes, with the strongest correlation between HF and LVESV_MRI_ (r_g_ = 0.31; p = 2.37 × 10^−11^; **FIGURE 1** and SUPPLEMENTAL TABLE 5).

**FIGURE 1:**
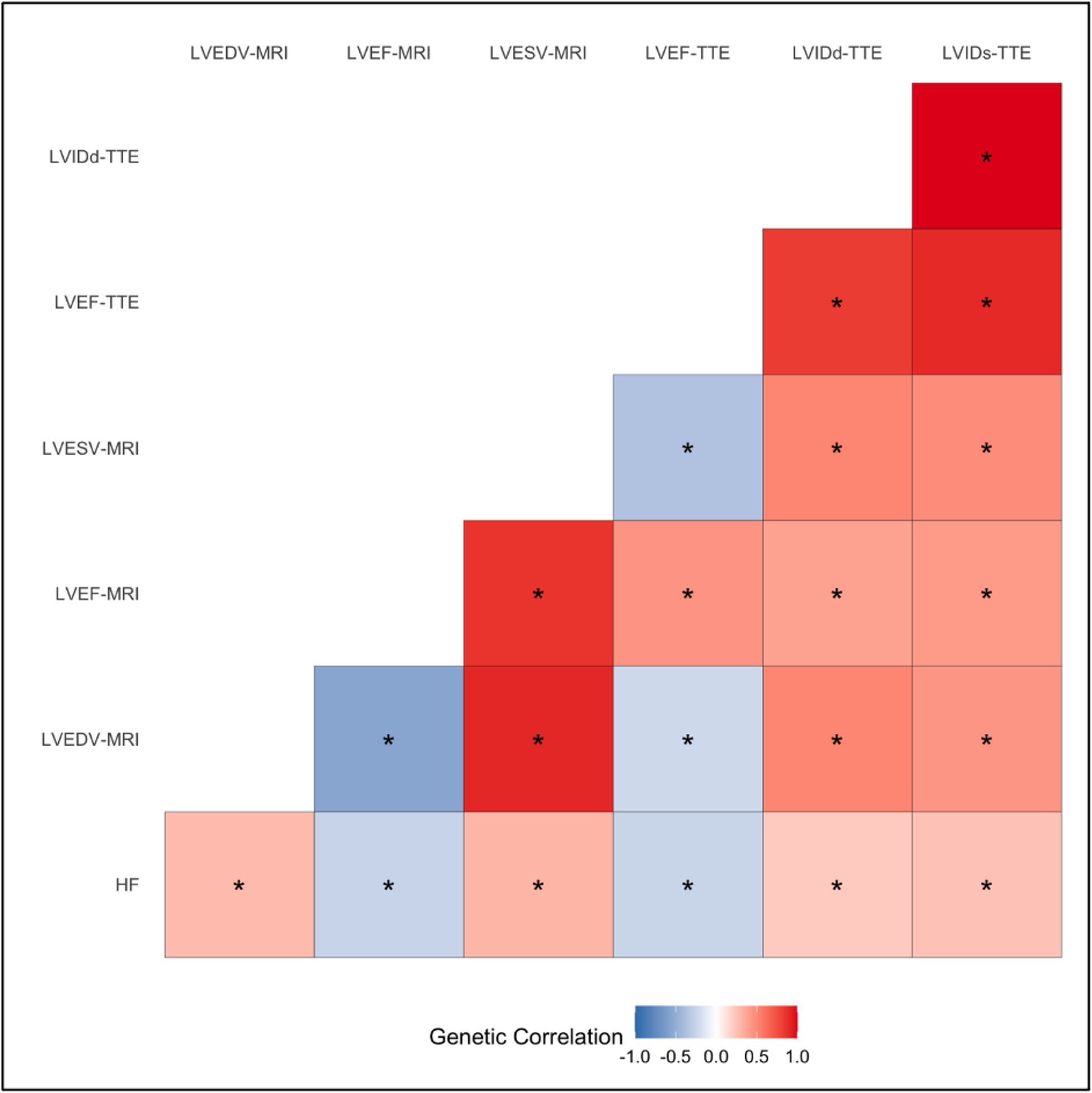
Genetic Correlation Among Heart Failure and Cardiac Imaging Traits. Cross-trait linkage disequilibrium score regression (LDSC) was performed using GWAS summary statistics for heart failure and cardiac imaging traits to estimate genetic correlation. Significant pairwise correlations after FDR adjustment for multiple testing (q < 0.05) are denoted with asterisks. MRI = magnetic resonance imaging; TTE = transthoracic echocardiogram; LVEDV = left ventricular end-diastolic volume; LVESV = left ventricular end-systolic volume; LVEF = left ventricular ejection fraction; LVIDd = left ventricular internal dimension in diastole; LVIDs = left ventricular internal dimension in systole.

### Multi-ancestry multivariate genome-wide analysis of HF endophenotypes identifies 16 novel loci

Having established significant genetic correlations between HF and cardiac imaging phenotypes suggestive of shared genetic etiology, we performed multivariate GWAMA to improve the power to discover associated genetic variants^9^. This method jointly considers related traits to estimate associations between genetic variants and a meta-trait, and is robust to scenarios where SNP-trait associations are derived from overlapping samples. This analysis utilized the results of the multi-ancestry HF meta-analysis performed above, along with the previously-published cardiac MRI^15^ and TTE^16^ GWAS to consider a composite HF “meta-trait”.

We identified 71 conditionally-independent variants (at 48 loci) associated with the composite heart failure meta-trait at GWS (**FIGURE 2** and SUPPLEMENTAL TABLES 6-7). Regional association plots at the novel loci demonstrated consistent genetic signals across multiple traits (**SUPPLEMENTAL FIGURE 3**). Of the 71 conditionally-independent variants, 37 had been previously reported and were significantly associated with multiple HF/imaging traits (**SUPPLEMENTAL TABLE 7**). The 34 remaining novel variants did not reach genome-wide significance in any of the parent studies (**SUPPLEMENTAL FIGURE 4A**). Among the 34 novel conditionally-independent variants, we identified 16 novel loci, where the lead variant was located >500kb away from any previously-reported genome-wide significant variant (**SUPPLEMENTAL FIGURE 4B**). Overall, lead variants at 12 of the 48 GWS loci were located within +/- 500kb of known cardiomyopathy genes, representing significant enrichment compared to matched control loci (one-tailed permutation p < 1 × 10^−4^) (**SUPPLEMENTAL FIGURE 5**). We did not detect enrichment of targets of approved medications indicated for treatment of heart failure or cardiomyopathy (p = 0.25).

**FIGURE 2:**
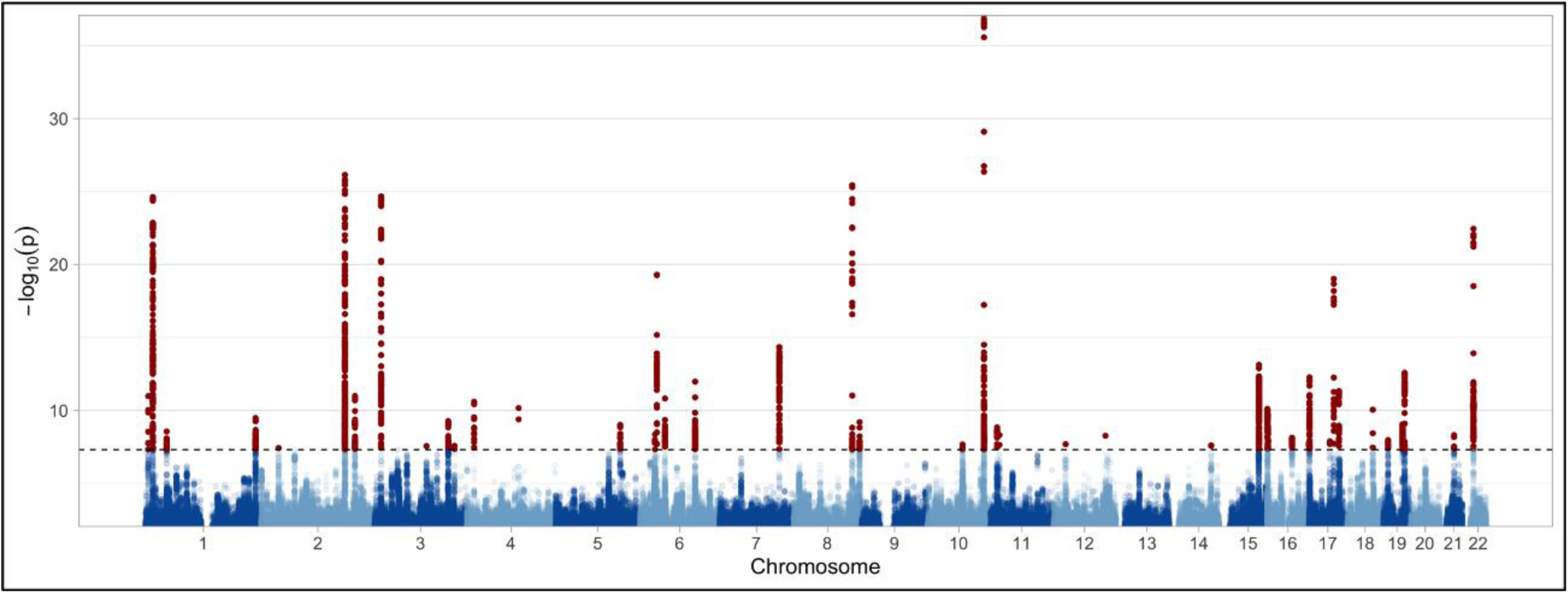
Results of Multivariate Genome Wide Association Study. Multivariate NGWAMA was performed to identify genetic variants associated with HF and cardiac structure/function traits. Manhattan plot depicting results of the multivariate analysis. Red dots denote variants meeting genome-wide significance (p < 5 × 10^−8^), marked by the dashed horizontal line.

To further assess the relevance of these loci across HF and imaging traits, we performed multi-trait colocalization^17^. We found evidence for colocalization across two or more HF/imaging traits at 40 of the 48 loci represented by the 71 conditionally independent variants, suggesting the multivariate GWAMA results represent discovery of shared genetic etiologies among the input traits (**SUPPLEMENTAL TABLE 8**).

The strongest novel association with the HF meta-trait was rs12921187, an intronic variant located within the *PPL* gene, which encodes periplakin, a cytoskeletal cross-linker protein. Mutations in desmoplakin, another member of this protein family, have been implicated in a form of arrhythmogenic cardiomyopathy associated with left ventricular fibrosis and systolic dysfunction^18^. The next strongest novel association was rs10113238, a variant located upstream of *FBXO32*, a muscle-specific F-box protein. This gene encodes a ubiquitin ligase that enables skeletal and cardiac muscle atrophy^19,20^, and has been implicated in familial dilated cardiomyopathy. Other strong novel associations include rs11079650, an intronic variant located within the *PRKCA* gene on chromosome 17. Functional studies of rs9912468, a *cis-*eQTL for *PRKCA* in perfect linkage disequilibrium with rs11079650 (EUR r^2^ = 1), previously identified an association in this region with expression of *PRKCA* in the human left ventricle, with zebrafish and in-vitro reporter assays suggestive of a cardiac-specific enhancer activity at this locus^21^. In humans, the *PRCKA* locus has previously been associated with electrocardiographic measures of left ventricular mass at genome-wide significance^21^, and nominally associated with echocardiographic traits and dilated cardiomyopathy^21^, now reaching genome-wide significance. Another strong novel association was rs56326533, an upstream variant located near the *SPATS2L* gene, which has previously been implicated in atrial fibrillation, QT-interval, and bronchodilator response in asthma^22^. Many of the novel loci in the multivariate analysis have been previously associated with known HF risk factors, including blood pressure, arrhythmias (eg. atrial fibrillation), and markers of inflammation (eg. circulating C-reactive protein levels), as well as anthropometric traits such as height and body fat/mass-related traits (**SUPPLEMENTAL TABLE 9**).

### Tissue and cell-type enrichment

To determine whether genetic associations with the composite HF meta-trait were enriched for specific tissues or cell-types we applied LDSC-SEG, a form of stratified of LD score regression which partitions heritability among sets of specifically expressed genes^23^. We detected significant associations (FDR < 0.05) with gene expression (GTEx) and chromatin marks (ROADMAP and ENTEX) in cardiac (eg. left ventricle, atrial appendage, aorta) and musculoskeletal tissue types (eg. quadriceps, psoas), as well as smooth-muscle found in the gastrointestinal system (eg. esophagus, stomach, colon) (**FIGURE 3**A-B and SUPPLEMENTAL TABLE 10). We leveraged single nucleus RNA sequencing (snRNA-seq) data from MAGNet to identify associations with cardiac-specific cell types, finding enrichment with cardiomyocytes (**FIGURE 3**C)^24^. Although many cardiometabolic traits are known to influence risk of HF, we did not detect significant enrichment of other tissue groups like adipose, blood, endocrine, or liver. While associations with cardiac and skeletal muscle were expected based on their mechanical role in maintaining normal cardiac structure/function, the absence of associations with classical metabolically-active tissues and cell-types may suggest that cardiac/musculoskeletal tissues and cell-types serve important metabolic roles in the pathogenesis of HF as well.

**FIGURE 3:**
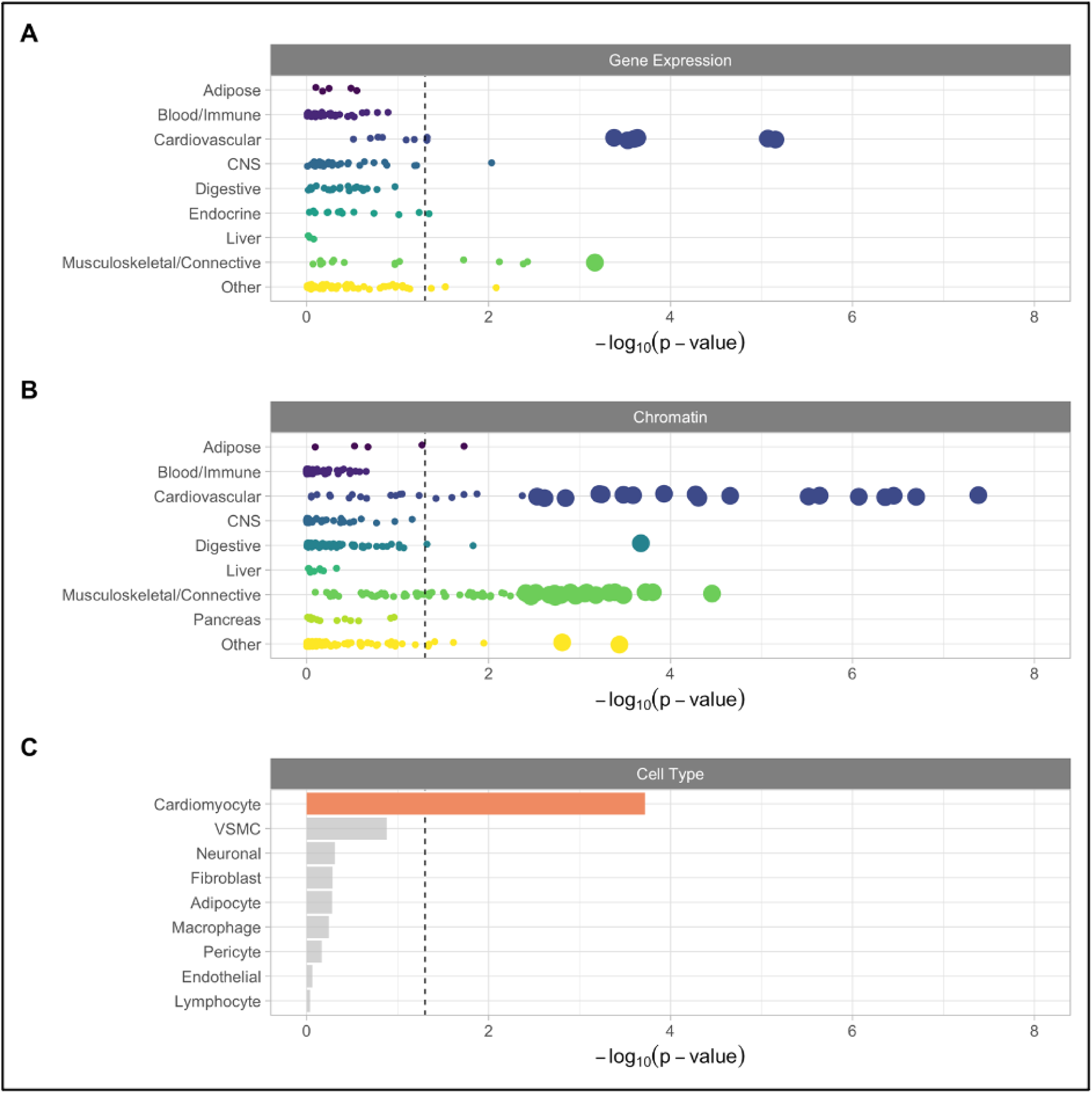
Tissue and Cell-type Associations. LDSC-SEG was performed to identify tissue and cell-type specific associations with the composite HF endophenotype. A) Association between tissue-specific gene expression (GTEx) and HF. B) Association between tissue-specific chromatin marks (ROADMAP and ENTEX). Large circles in panels A and B represent significant (FDR q < 0.05) associations. C) Associations with cardiac-specific cell-types based on differential gene expression. Colored bar denotes FDR < 0.05.

### Colocalization, transcriptome-wide association, and gene-expression profiling analyses prioritize HF effector genes

#### Multi-Trait Colocalization

To prioritize putative candidate genes associated with HF risk, we sought several lines of evidence. We first utilized an expression quantitative trait loci (eQTL) dataset from the Myocardial Applied Genomics Network (MAGNet), derived from 313 human hearts (177 failing hearts, 136 donor nonfailing control hearts)^25^. We performed multi-trait colocalization to identify shared genetic signals associated with gene expression in the heart and HF, MRI, and TTE traits. In total, expression of 21 genes in healthy and/or failing hearts colocalized with HF and/or cardiac imaging traits (TABLE 2: Multi-trait Colocalization of Gene Expression, HF, and Quantitative Cardiac Imaging traits**TABLE 2**). The set of genes with evidence for colocalization across the largest number of gene expression, HF, and imaging traits included *MMP11, SLC2A11, PRKCA*, and *SPON1*, which colocalized with 9, 8, 7, and 6 HF endophenotypes, respectively (TABLE 2: Multi-trait Colocalization of Gene Expression, HF, and Quantitative Cardiac Imaging traits**TABLE 2**).

**TABLE 2:** Multi-trait Colocalization of Gene Expression, HF, and Quantitative Cardiac Imaging traits.

| Gene | Traits | Posterior Probability | Regional Probability | Candidate SNP | SNP Posterior |
| --- | --- | --- | --- | --- | --- |
| DNAJC18 | Healthy, Diseased, LVEF-MRI, LVESV-MRI | 0.9899 | 0.9993 | rs11242465 | 1.0000 |
| BCKDHA | Healthy, Diseased, LVEDV-MRI, LVESV-MRI | 0.9755 | 0.9996 | rs3810175 | 0.9778 |
| DUSP13 | Healthy, LVEF-MRI, LVESV-MRI | 0.9744 | 0.9969 | rs4745763 | 0.9973 |
| ALPK3 | Diseased, LVEF-MRI, LVEDV-MRI, LVESV-MRI | 0.9679 | 0.9957 | rs17599989 | 0.5535 |
| LMF1 | Healthy, Diseased, LVEF-MRI, LVESV-MRI | 0.9513 | 0.9956 | rs4984621 | 0.7122 |
| EXOSC5 | Healthy, Diseased, LVEDV-MRI, LVESV-MRI | 0.9498 | 0.9822 | rs4803465 | 0.7151 |
| UBE2Q2P1 | Diseased, LVEF-MRI, LVEDV-MRI, LVESV-MRI | 0.8923 | 0.9434 | rs17599989 | 0.6692 |
| MLF1 | Healthy, Diseased, LVEF-MRI, LVESV-MRI | 0.8059 | 0.8337 | rs891464 | 0.8290 |
| SSTR5 | Diseased, LVEF-MRI, LVESV-MRI | 0.7562 | 0.8097 | rs4984621 | 0.4276 |
| FOXN3 | Healthy, Diseased, LVEF-MRI, LVEDV-MRI, LVESV-MRI | 0.7391 | 0.7576 | rs1952183 | 0.9848 |
| HSPB7 | Diseased, CHF, LVEF-MRI, LVEDV-MRI, LVESV-MRI | 0.7186 | 0.7350 | rs1763610 | 1.0000 |
| SPON1 | Healthy, Diseased, LVEF-MRI, LVEDV-MRI, LVESV-MRI, LVEF-TTE | 0.6757 | 0.7258 | rs10832164 | 0.9118 |
| INPP5K | Diseased, LVEF-MRI, LVEDV-MRI, LVESV-MRI | 0.6153 | 0.6581 | rs15948 | 0.9999 |
| PRKCA | Healthy, Diseased, LVEF-MRI, LVEDV-MRI, LVESV-MRI, | 0.6101 | 0.6238 | rs9912468 | 0.5134 |
|  | LVIDd-TTE, LVIDs-TTE |  |  |  |  |
| MLST8 | Diseased, LVEDV-MRI, LVESV-MRI | 0.5812 | 0.5828 | rs26835 | 1.0000 |
| MMP11 | Healthy, Diseased, CHF, LVEF-TTE, LVIDd-TTE, LVIDs-TTE, LVEF-MRI, LVEDV-MRI, LVESV-MRI | 0.5664 | 0.5755 | rs5760061 | 0.9226 |
| SEC23IP | Diseased, LVEF-TTE, LVIDs-TTE | 0.5474 | 0.5999 | rs17099199 | 0.6403 |
| COPB1 | Healthy, LVEF-MRI, LVEDV-MRI, LVESV-MRI, LVEF-TTE | 0.5413 | 0.6086 | rs1528640 | 0.5354 |
| MMACHC | Diseased, LVEF-MRI, LVEDV-MRI, LVESV-MRI | 0.5089 | 0.6421 | rs12743512 | 0.5021 |
| SLC2A11 | Healthy, CHF, LVEF-TTE, LVIDd-TTE, LVIDs-TTE, LVEF-MRI, LVEDV-MRI, LVESV-MRI | 0.4450 | 0.5916 | rs5760061 | 0.8185 |
| CDKN1A | Healthy, CHF, LVEF-MRI, LVESV-MRI | 0.2606 | 0.5113 | rs1321310 | 0.9938 |

Multi-trait colocalization was performed using HyPrColoc to integrate eQTL data from Healthy and Diseased hearts from the MAGNet consortium with GWAS of heart failure (multi-ancestry meta analysis; CHF), and cardiac imaging traits (MRI and TTE). MRI = magnetic resonance imaging; TTE = transthoracic echocardiogram; LVEDV = left ventricular end-diastolic volume; LVESV = left ventricular end-systolic volume; LVEF = left ventricular ejection fraction; LVIDd = left ventricular internal dimension in diastole; LVIDs = left ventricular internal dimension in systole.

Among genes with the strongest evidence for a shared genetic etiology between cardiac expression levels and the HF meta-trait was *BCKDHA. BCKDHA* encodes branched chain keto acid dehydrogenase E1 subunit alpha, a key enzyme responsible for branch chain amino acid (BCAA) degradation, a pathway previously implicated in adverse cardiac remodeling and heart failure^26–28^. We found strong evidence for colocalization between *BCKDHA* expression in healthy hearts, failing hearts, LVEDV_MRI_ and LVESV_MRI_ (posterior probability 0.98). *BCKDHA* expression was also increased in failing compared to healthy hearts (EUR fold change = 1.25, p = 0.005; AFR fold change = 1.24, p = 0.043). Given the colocalization evidence for a shared genetic risk factor influencing *BCKDHA* expression and LVEDV_MRI_ and LVESV_MRI_, we performed Mendelian randomization to determine whether circulating branch-chain amino acid (isoleucine, leucine, valine) levels may be causally associated with LVEDV_MRI_ and LVESV_MRI_. Using genetic instruments derived from a GWAS of circulating metabolites among up to 24,925 participants of ten European studies,^29^ Increased circulating levels of isoleucine and leucine were significantly associated with decreased LVEDV_MRI_ (leucine β = -0.137, 95% CI -0.25 to -0.022, p = 0.02; isoleucine β = -0.276, 95% CI -0.38 to -0.17, p = 3 × 10^−7^) and LVSEV_MRI_ (leucine β = -.131, 95% CI - 0.24 to -0.026, p = 0.01; isoleucine β = -0.217, 95% CI -0.33 to -0.11, p = 1 × 10^−4^), with no significant associations identified for valine (**FIGURE 4**). We did not detect evidence of reverse-causality (eg. increased LVEDV_MRI_ or LVESV_MRI_ leading to increased BCAA levels), and the findings remained robust when using the weighted-median MR method, which makes different assumptions about the presence of pleiotropy (SUPPLEMENTAL **FIGURE 5**).

**FIGURE 4:**
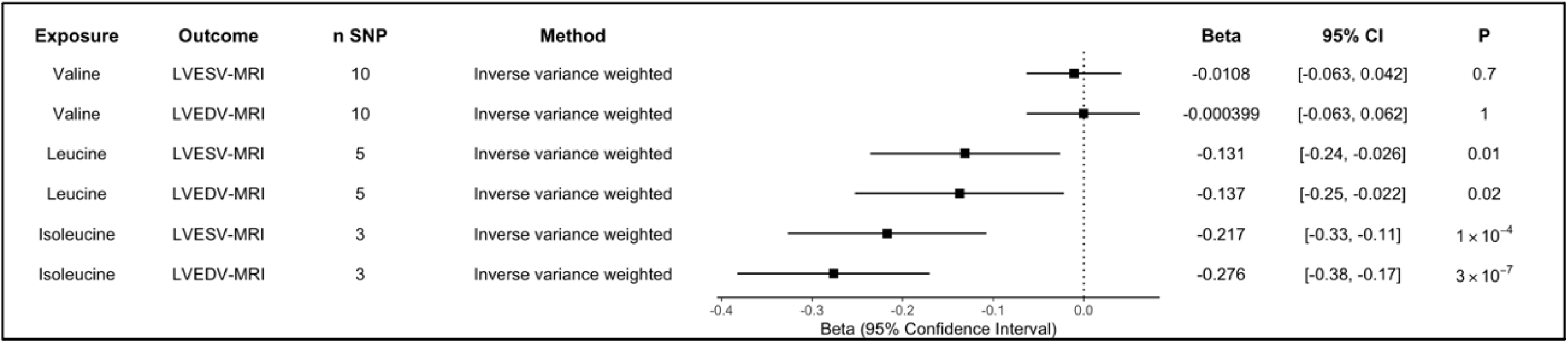
Branch Chain Amino Acid Mendelian Randomization. Mendelian randomization was performed to identify whether circulating branch chain amino acid levels were associated with cardiac MRI traits, given colocalization between BCKDHA and both LVESV_MRI_ and LVEDV_MRI_.

#### Transcriptome-Wide Association Study (TWAS)

Next, we performed transcriptome-wide association studies (TWAS) integrating gene expression and splicing data from the Genotype-Tissue Expression (GTEx) project with the results of our multivariate GWAMA study^30–33^. We performed TWAS using models from cardiometabolic tissues (left ventricle, atrial appendage, visceral adipose, subcutaneous adipose, liver, kidney, and blood), to identify genes where tissue-specific expression levels (eQTL) or transcript splicing events (sQTL) may be relevant to HF. Across all tissues, we identified 66 distinct genes representing 150 gene-tissue pairs where gene expression was significantly associated with HF endophenotypes after Bonferroni adjustment for multiple testing (75,563 gene-tissue pairs) (**FIGURE 5A**; **SUPPLEMENTAL TABLE 11**). This set of 66 genes was enriched for established cardiomyopathy genes (hypergeometric p = 0.007). We also identified 339 splicing events across 54 genes that were significantly associated with HF endophenotypes traits after Bonferroni adjustment for multiple testing (173,031 splicing event-tissue pairs) (**FIGURE 5B**; **SUPPLEMENTAL TABLE 12**). This set of 54 genes was also enriched for Mendelian cardiomyopathy genes (hypergeometric p = 0.011). Of the 54 significant splicing-associated genes, 26 were also identified in the eQTL TWAS. Several canonical cardiomyopathy genes were identified only in the sQTL analysis, including *BAG3* and *TTN*.

**FIGURE 5:**
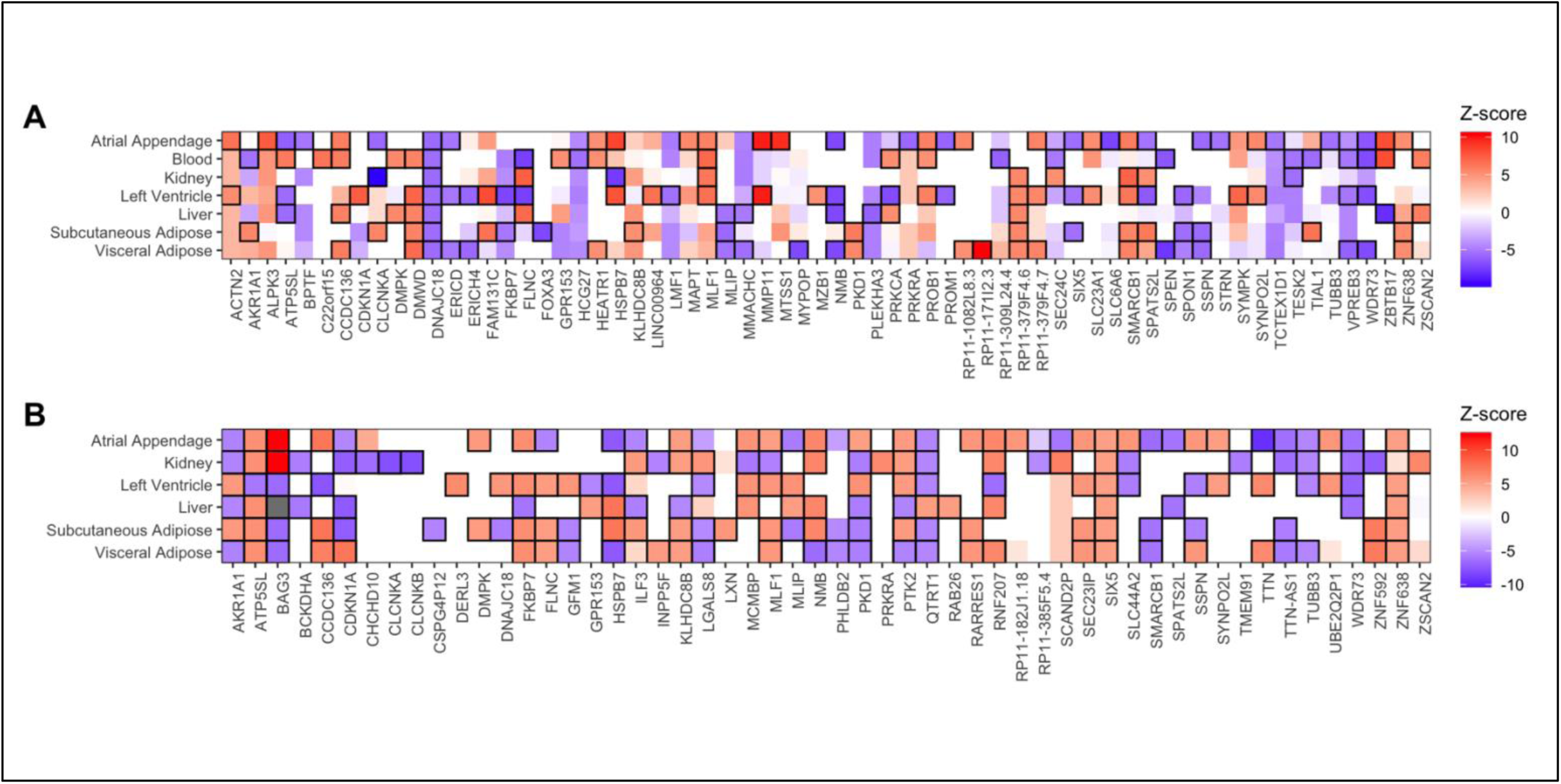
TWAS Results. TWAS identified 66 distinct genes (representing 150 gene-tissue pairs) where expression was associated with adverse HF/structure/function traits, and 54 distinct genes (across 339 splicing-tissue pairs) where splicing was associated with adverse HF/structure/function traits. A) Heatmap depicting the gene-tissue pairs where gene expression was significantly associated with adverse HF/structure/function traits. B) Heatmap depicting the gene-tissue pairs where transcript splicing was significantly associated with adverse HF/structure/function traits. Black borders surround gene-tissue associations that were significant (p < 0.05) after Bonferroni adjustment for multiple testing.

Among the most highly prioritized associations was *CLCNKA* gene expression in kidney (p = 2.80 × 10^−23^ in eQTL and p = 2.35 × 10^−25^ in sQTL analyses). *CLCNKA* encodes the K_a_ renal chloride channel (ClC-K_a_), with a prior candidate-variant study identifying a suggestive association between the common coding variant rs10927887 and heart failure^34^. Further functional characterization of this variant revealed loss-of-function in the ClC-K_a_ chloride channel, implicating a Bartter syndrome-like cardio-renal axis in heart failure^34^. Although other genes in this region including *HSPB7*^35^ and *ZBTB17*^36^ have been implicated in heart failure, our results provide support for a role of *CLCNKA. MMP11*, which encodes Matrix metalloproteinase 11, an enzyme responsible for degradation of several extracellular matrix substrates, was also highly prioritized (expression in left ventricle, p = 1.24 × 10^−22^; expression in atrial appendage, p = 9.55 × 10^−23^). Matrix metalloproteinase inhibition has been associated with favorable echocardiographic measures of remodeling following myocardial infarction in animal studies^37^. Expression of *MTSS1* in atrial appendage tissue was also highly prioritized (p = 1.10 × 10^−20^). *MTSS1* encodes Metastasis suppressor protein 1. Candidate-variant studies of this locus have identified significant associations between the lead variant at this locus (rs12541595) and heart failure-related traits in humans, and knockout of this gene in mice has been associated with changes in echocardiographic measures related to heart failure^38^.

#### Gene Expression Profiling

To validate the eQTL TWAS findings we compared expression levels of genes prioritized by the TWAS analyses (Bonferroni p < 0.05) among 166 healthy (122 EUR, 44 AFR) and 166 failing (89 EUR, 77 AFR) hearts from MAGNet. Of 57 genes with available expression data, we identified 40 genes where expression significantly differed between healthy and failing hearts after Bonferroni adjustment for multiple testing (p < 0.05 adjusting for 57 genes) (**FIGURE 6**; SUPPLEMENTAL TABLE 13).

**FIGURE 6:**
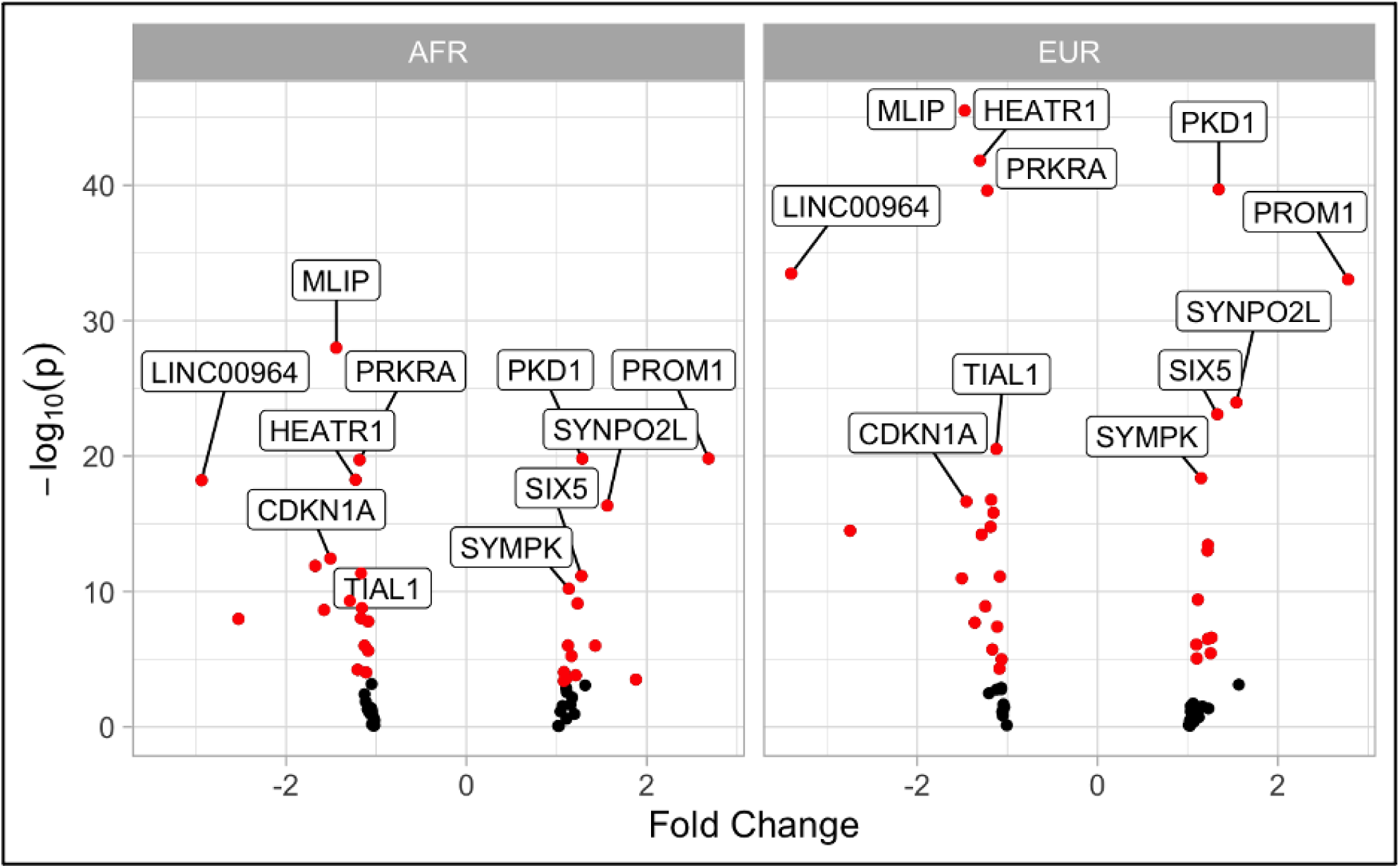
Gene Expression Profiling Results. Left ventricular gene expression profiling from MAGNet for genes prioritized by TWAS. Red dots represent candidate genes with significant differential expression among failing vs. healthy hearts, after Bonferroni adjustment for multiple testing. The genes with the most significant differential expression between healthy and failing hearts are labeled.

#### Gene Ontology

We performed an exploratory gene-ontology analysis^39^ among a broader set of 334 eQTL TWAS-prioritized genes (FDR < 0.05). Cellular Component analysis was notable for significant enrichment (FDR < 0.05) of cardiomyocyte components including Z-disc, I-band, myofibril, sarcomere, and sarcoplasm, reinforcing the importance of these cellular substructures in the pathogenesis of HF (**SUPPLEMENTAL TABLE 14**).

### Proteome-wide Mendelian randomization prioritizes circulating proteins associated with adverse HF phenotypes

Finally, we performed an unbiased proteome-wide Mendelian randomization analysis using high-confidence genetic instruments for 725 circulating proteins to identify their contribution to each of the HF endophenotypes. We identified 18 significant (FDR < 0.05) protein-trait associations, across 11 distinct circulating proteins (**FIGURE 7**A). Among these significant gene-trait pairs was lipoprotein(a) (*LPA*), with increasing levels associated with increased risk of HF (OR 1.09 per 1-SD increase in lipoprotein(a), 95% CI 1.06 to 1.11, p = 1.6 × 10^−11^). Lipoprotein(a) is a known risk factor for coronary artery disease that has been previously associated in observational studies with incident heart failure and associated hospitalization^40,41^. To further evaluate this association, we considered an additional genetic instrument consisting of 3 genetic variants previously reported to explain >40% of the variation in circulating Lp(a) levels across multiple cohorts^42^. Using this genetic instrument, each 10mg/dL increase in Lp(a) was associated with increased risk of heart failure (OR 1.03, 95% CI 1.021 to 1.031, p = 1.95 × 10^−21^). This association was not attenuated in multivariable MR accounting for the association of these genetic variants with coronary artery disease (OR 1.03, 95% CI 1.022 to 1.038, p = 1.72 × 10^−13^), indicating a potentially independent role of lipoprotein(a) in the pathogenesis of HF.

**FIGURE 7:**
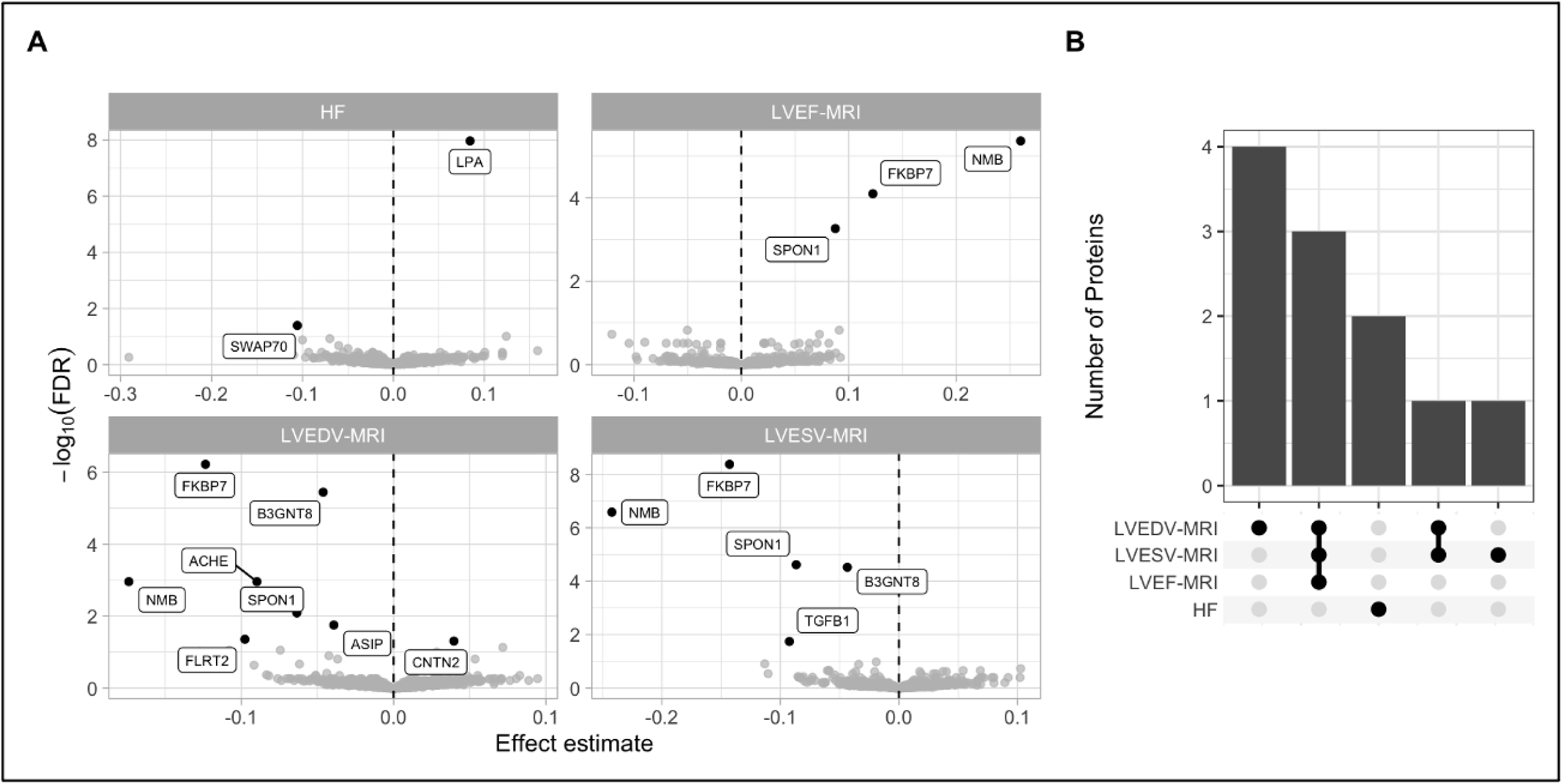
Proteome-wide Mendelian Randomization. Proteome-wide MR was performed using high-confidence genetic instruments to detect associations between circulating proteins and cardiac endophenotypes. A) Associations between circulating protein levels and HF traits. B) Number of shared associations across HF traits.

We additionally identified 4 circulating proteins that were associated with multiple traits (**FIGURE 7**B), including *SPON1, NMB*, and *FKBP7*, which were each associated with increased LVEF and decreased ventricular volumes, and *B3GNT8* which was associated with decreased volumes (**FIGURE 7**A). Circulating *SPON1* and *FKBP7* have both been previously linked to atrial fibrillation, a known HF risk factor^43^, and *SPON1* has been identified as a potential biomarker for HF hospitalization^44^.

## DISCUSSION

In this study we performed multi-ancestry and multi-trait genome-wide meta-analyses of HF and related cardiac imaging traits. We analyzed the genetic relationships among these traits 1) demonstrating improved power for novel locus discovery for this collection of traits, 2) implicating both known and novel variants in HF pathogenesis and 3) identifying circulating proteins that may represent therapeutic targets for HF.

First, these findings highlight the value of multi-ancestry and multi-trait genome wide analyses to improve genetic discovery. Although GWAS of HF have been performed largely in populations of European ancestry, HF is a global disease associated with high morbidity and mortality. Consistent with GWAS of other cardiovascular traits like CAD^8^, we found that multi-ancestry analysis improved power for discovery, identifying 4 novel all-cause HF loci that had previously been associated with cardiometabolic and anthropometric traits that are themselves HF risk factors. Similarly, incorporating HF endophenotypes in multi-trait analyses further improved power for discovery. Interestingly, while the cardiac MRI GWAS was largely restricted to healthy individuals, our genetic correlation analysis found that the genetic architecture of phenotypic variation within the normal range was strongly linked to pathologic HF. Leveraging this relationship, we found that jointly considering HF with closely-related imaging endophenotypes was associated with a substantial 29% improvement in locus discovery. With the growth of institutional biobanks that link rich electronic health records, laboratory, and imaging findings with genetic data, more refined phenotyping efforts (including studies focused on variation within the normal range among otherwise healthy individuals) may further enable genetic discovery within this multi-trait paradigm.

Second, we found that loci from the multivariate GWAS implicated known and novel HF effector genes and pathways. For example, we detected significant enrichment of known Mendelian cardiomyopathy-associated genes in both our NGWAMA and TWAS analyses, with TWAS results further enriched for genes associated with cellular contractile machinery. Lead NGWAMA variants were also associated with common cardiometabolic and anthropometric risk factors for HF. We additionally found convergent evidence for the influence of common variants across several secondary analyses. For example, *CLCNKA* was associated with HF across several lines of evidence, implicated in both multivariate GWAS and eQTL TWAS, and sQTL TWAS analyses. MMP11 was also identified across several analyses, including multivariate GWAS, TWAS, multi-trait colocalization, and gene-expression profiling of healthy and failing hearts. Overall, these findings highlight the utility of integrative analyses that draw on genetic association, gene expression, chromatin modification, and tissue/cell-type specific datasets to prioritize disease-associated genes. In sum, these results suggest that genes, tissues, and cellular components associated with Mendelian forms of HF are important for common manifestations of HF as well.

Finally, we observed potentially causal links between circulating metabolites and proteins with HF and related imaging traits. Elevated circulating levels of branch chain amino acids have been previously implicated as a risk factor for incident HF and adverse cardiac remodeling in a murine model of myocardial infarction^45,46^. Our colocalization and MR analyses identified strong evidence of a shared genetic etiology and potentially causal relationship between circulating leucine and isoleucine levels and left ventricular volumes among healthy individuals, although the pathologic relevance to human HF specifically requires further investigation. Our proteome-wide MR analysis similarly identified several proteins associated with HF and cardiac imaging traits, including SPON1, which was also prioritized in our multi-trait colocalization, TWAS, and gene-expression analyses.

### Limitations

This study has several possible limitations. First, although we performed multi-ancestry GWAS and multivariate GWAMA, these analyses focused primarily on participants of European and Japanese ancestry. As the global burden of heart failure is increasing, future GWAS of HF and related traits in other diverse populations is warranted. Future multi-ancestry analyses will hopefully further improve our understanding of the true breadth of the common genetic basis of these traits. In downstream colocalization, TWAS, and gene expression profiling analyses we included multi-ancestry cohorts (GTEx v8, MAGNet), which should overall improve generalizability of our findings. Second, interpreting the absolute effect estimates of the multivariate GWAMA analysis can be challenging as they reflect a composite trait. We included highly-correlated imaging traits with biologically plausible connection to HF maximize interpretability, however including other cardiovascular and HF endophenotypes may further improve discovery. Third, these results represent the findings of a single multi-trait GWAS method, and whether locus discovery or biologic relevance may be further improved with other methods requires further study. Our GWAS and GWAMA results consider only common genetic variants imputed to the 1000 Genomes Project phase 3 reference panel, and whether additional discovery is enabled by recently released state-of-the art imputation panels like TOPMed will require further study.

## Conclusion

In summary, these analyses highlight similarities and differences among heart failure and associated cardiovascular imaging endophenotypes, implicate novel common genetic variation in the pathogenesis of HF, and identify circulating proteins that may represent novel cardiomyopathy treatment targets.

## METHODS

### HF Meta-analysis

GWAS summary statistics for heart failure were obtained from HERMES (N = 47,309 cases and 930,014 controls; http://kp4cd.org/datasets/mi) and BioBank Japan (N = 9,413 cases and 203,040 controls; http://jenger.riken.jp/en/result). Both studies included individuals with all-cause heart failure. Details of genotyping and phenotyping for each study have been previously reported^15,47^. Fixed-effects meta-analysis was performed using METAL^48^ using the inverse-variance weighted (standard error) method for the set of variants with MAF > 0.01 present in the 1000 Genomes Phase 3 reference panel. Independent significant SNPs, lead SNPs, and genomic risk loci were defined locally using PLINK^49^ and the 1000 Genomes Phase 3 reference panel, based on the default r^2^ and distance thresholds utilized by the online FUMA tool^50^. We employed the conventional genome-wide significance threshold of p < 5 × 10^−8^ to characterize significant associations. Replication for significant loci was performed using publicly available HF GWAS data from Release 4 of FinnGen (http://r4.finngen.fi/pheno/I9_HEARTFAIL_ALLCAUSE).

### Genetic Correlation of HF and Cardiac Imaging Traits

Cross-trait linkage-disequilibrium score regression (LDSC) was performed to estimate genetic correlation (rg) between HF, cardiac MRI (LVEF, LVEDV, LVESV; UKB http://kp4cd.org/datasets/mi), and echocardiographic (LVEF, LVIDD, LVIDS; BBJ http://jenger.riken.jp/en/result) traits^13,14^. LDSC is a computationally-efficient method which utilizes GWAS summary statistics to estimate heritability and genetic correlation between polygenic traits.

### Multivariate GWAMA

Genome-wide association study summary statistics were obtained for heart failure (HERMES+BBJ), cardiac MRI (LVEF, LVEDV, LVESV), and echocardiographic (LVEF, LVIDD, LVIDS) traits. Variants were filtered to include common (MAF > 0.01) variants present in the 1000 Genomes Phase 3 reference panel. Cross-trait linkage-disequilibrium score regression (LDSC) was performed to estimate genetic correlation (rg)^13,14^. Cross-trait LD-score intercepts were estimated to quantify the dependence between traits and account for sample overlap. N-weighted Multivariate GWAMA was performed to estimate multivariate SNP effects for the composite HF endophenotype^9^. As recommended, the effect estimates for MRI-LVEF and TTE-LVEF were reversed to ensure positive correlation with the other traits.

### Cardiomyopathy Gene Enrichment

We utilized a previously published list of Mendelian cardiomyopathy-associated genes^15^ aggregated from commercially-available gene panels to test for enrichment of GWAMA loci. In sum, these panels contained 108 autosomal genes. To test enrichment of GWAMA loci we used SNPsnap to identify 10,000 matched SNPs from across the genome for each lead GWAMA SNP based on allele frequency, number of SNPs in LD, distance to nearest gene, and gene density^51^. We compared the number of GWAMA variants located within 500kb of Mendelian cardiomyopathy-associated genes to the null baseline established across 10,000 permutations of matched SNPs to yield an empirical one-tailed p-value.

### Multi-trait colocalization

Statistical colocalization is a method to assess shared genetic etiology between traits. We used HyPrColoc^17^, a recently developed Bayesian algorithm designed to simultaneously and efficiently evaluate for colocalization across multiple traits using summary statistics. We assessed for colocalization across HF, MRI, TTE, and heart gene expression traits in the 500kb region centered on the lead variants identified in the multivariate GWAMA analysis. Gene expression data was derived from an expression quantitative trait loci (eQTL) dataset from the Myocardial Applied Genomics Network (MAGNet), derived from 313 human hearts (177 failing hearts, 136 donor nonfailing control hearts) obtained at time of organ procurement (control hearts) or heart transplant (failing hearts)^25^. Evidence for colocalization was determined based on the default variant specific regional and alignment priors 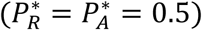, with colocalization identified when *P*_*R*_*P*_*A*_ ≥ 0.25.

### Tissue and cell-type enrichment

We implemented LDSC-SEG^23^ to test for enrichment of disease heritability by integrating our GWAMA summary statistics with gene expression^52,53^, chromatin^54,55^, and cardiac-specific cell-type^24^ datasets. We applied false discovery rate correction separately across each analysis (gene expression, chromatin, and cardiac cell-type) to account for multiple testing, with FDR < 0.05 considered significant.

### Branched Chain Amino Acid Mendelian Randomization

Genetic variants associated with branch chain amino acids (leucine, isoleucine, and valine) at genome wide significance (p < 5 × 10^−8^) were identified from a study including up to 24,925 participants of ten European studies who underwent NMR profiling of circulating metabolites^29^. Genetic instruments were constructed from independent (EUR r^2^ < 0.3, distance = 10,000kb) variants associated with each BCAA at genome-wide significance. The corresponding SNP effects were identified in GWAS summary statistics for LVEDV_MRI_ and LVSEV_MRI_, harmonized to consistent effect alleles, and two-sample inverse variance weighted Mendelian randomization with random effects was performed using the *TwoSampleMR* package in R^56^. Sensitivity analysis was performed using the weighted median method, which remains robust when up to 50% of the weight of the genetic instrument is invalid^57^.

### Transcriptome-wide Association Study

S-PrediXcan was used to integrate gene expression and splicing data from GTEx version 8 and GWAS summary statistics from the multivariate GWAMA experiment to identify genes associated with adverse HF phenotypes^30–33^. Pretrained gene expression and transcript splicing models from cardiometabolic tissues (left ventricle, atrial appendage, visceral adipose, subcutaneous adipose, liver, and kidney) were obtained from http://predictdb.org/. Bonferroni adjustment was performed to account for multiple testing (75,563 gene-tissue pairs for eQTL TWAS; 173,031 splicing event-tissue pairs for sQTL TWAS), with p < 0.05 considered significant. We tested for enrichment of TWAS-prioritized genes among Mendelian cardiomyopathy-associated genes using the hypergeometric distribution to yield a 1-tailed p-value.

### Cardiac Gene Expression Profiling

The Myocardial Applied Genetics Consortium (MAGnet) is a multicenter, institutional review board approved consortium designed to explore the genetic underpinnings of cardiac gene expression^25,58^. Briefly, human cardiac samples were obtained from failing hearts collected at time of heart transplantation, and healthy donor hearts that were suitable for transplant but logistically did not reach recipients. RNA gene expression profiling on cardiac tissue samples obtained from the left ventricle was performed as previously described^25^. Gene expression data was queried to determine whether significant expression differences existed between healthy and failing hearts for genes prioritized by TWAS, with significant fold-changes differences determined by Bonferroni-adjusted p < 0.05 to account for multiple testing.

### Cellular Component Analysis

Cellular component analysis was performed using ShinyGO, an online platform for gene enrichment analysis^39^. Based on a set of input genes, the application tests for enrichment among prespecified gene sets based on the hypergeometric distribution followed by false discovery rate correction for multiple testing. Components with FDR q < 0.05 were considered significant.

### Proteome-Wide Mendelian Randomization

Proteome-wide Mendelian Randomization was performed as previously described (http://www.epigraphdb.org/pqtl/)^59^. Briefly, we identified high-confidence (tier 1) *cis*-acting genetic instruments for 725 circulating proteins, which passed previously defined consistency and pleiotropy tests and had available corresponding SNP effects from our HF meta-analysis and/or the cardiac MRI GWAS. When multiple SNPs were available for an exposure-outcome pair, inverse-variance weighted MR was performed as the primary analysis, with Wald-ratio MR performed when only one SNP was available for the exposure-outcome pair. FDR correction was applied to account for multiple testing, with q < 0.05 considered significant. All statistical analyses were performed using R version 4.0.3 (R Foundation for Statistical Computing, Vienna, Austria).

## Data Availability

GWAS summary statistics for heart failure (HERMES: http://kp4cd.org/datasets/mi; BioBank Japan: http://jenger.riken.jp/en/result; FinnGen: http://r4.finngen.fi/pheno/I9_HEARTFAIL_ALLCAUSE), cardiac MRI (http://kp4cd.org/datasets/mi, and echocardiography traits (http://jenger.riken.jp/en/result) are publicly available. Cardiac eQTL and RNA expression/sequencing data were provided by the Myocardial Applied Genomics Network (MAGNet).

## ETHICAL APPROVAL

The UK Biobank obtained IRB approval from the North West Multi-centre Research Ethics Committee (approval number: 11/NW/0382), and participants provided informed consent. The BioBank Japan Project was approved by the research ethics committees at the Institute of Medical Science, the University of Tokyo, the RIKEN Yokohama Institute, and cooperating hospitals; participants gave written informed consent. FinnGen participants provided informed consent for biobank research, and the Coordinating Ethics Committee of the Hospital District of Helsinki and Uusimaa (HUS) approved the FinnGen Study protocol No. HUS/990/2017.

