## Supplemental Figures for "Multi-ancestry Multivariate Genome-Wide Analysis Highlights the Role of Common Genetic Variation in Cardiac Structure, Function, and Heart Failure-related Traits"

**SUPPLEMENTAL MATERIAL**


**SUPPLEMENTAL FIGURES 2**

SUPPLEMENTAL FIGURE 1: HEART FAILURE GENOME WIDE ASSOCIATION META-ANALYSIS MANHATTAN PLOT 3
SUPPLEMENTAL FIGURE 2: HEART FAILURE GENOME WIDE ASSOCIATION META-ANALYSIS QUANTILE-QUANTILE PLOT 4
SUPPLEMENTAL FIGURE 3: N-GWAMA REGIONAL ASSOCIATION PLOTS 5
SUPPLEMENTAL FIGURE 4: OVERLAP OF GENOME-WIDE SIGNIFICANT VARIANTS ACROSS GWAS STUDIES 14
SUPPLEMENTAL FIGURE 5: ENRICHMENT OF CARDIOMYOPATHY GENES IN GWAMA LOCI 15
SUPPLEMENTAL FIGURE 6: BRANCH CHAIN AMINO ACID WEIGHTED MEDIAN MENDELIAN RANDOMIZATION 16

### SUPPLEMENTAL FIGURES

**Supplemental Figure 1: Heart Failure Genome Wide Association Meta-Analysis Manhattan Plot**

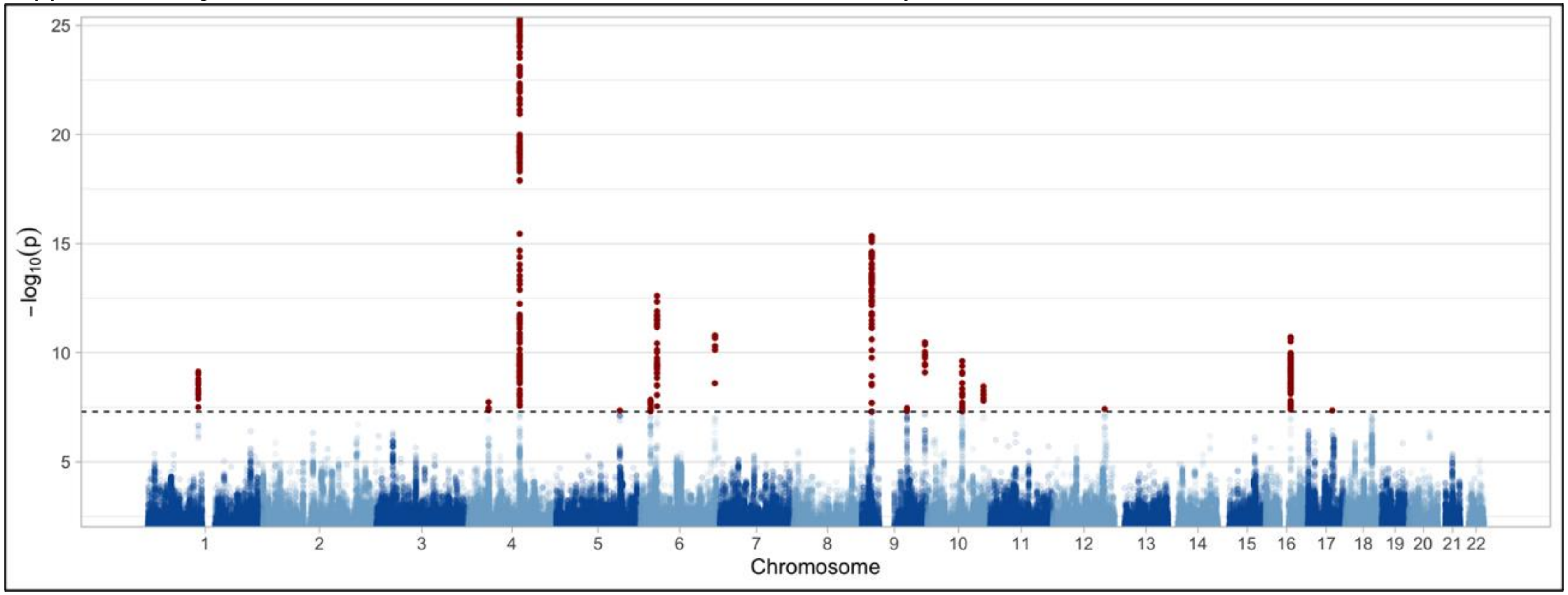


Manhattan plot demonstrating association between genetic variants and heart failure. Red points denote variants that exceed the genome-wide significance threshold ($p < 5 \times 10^{-8}$).

**Supplemental Figure 2: Heart Failure Genome Wide Association Meta-Analysis Quantile-Quantile Plot**

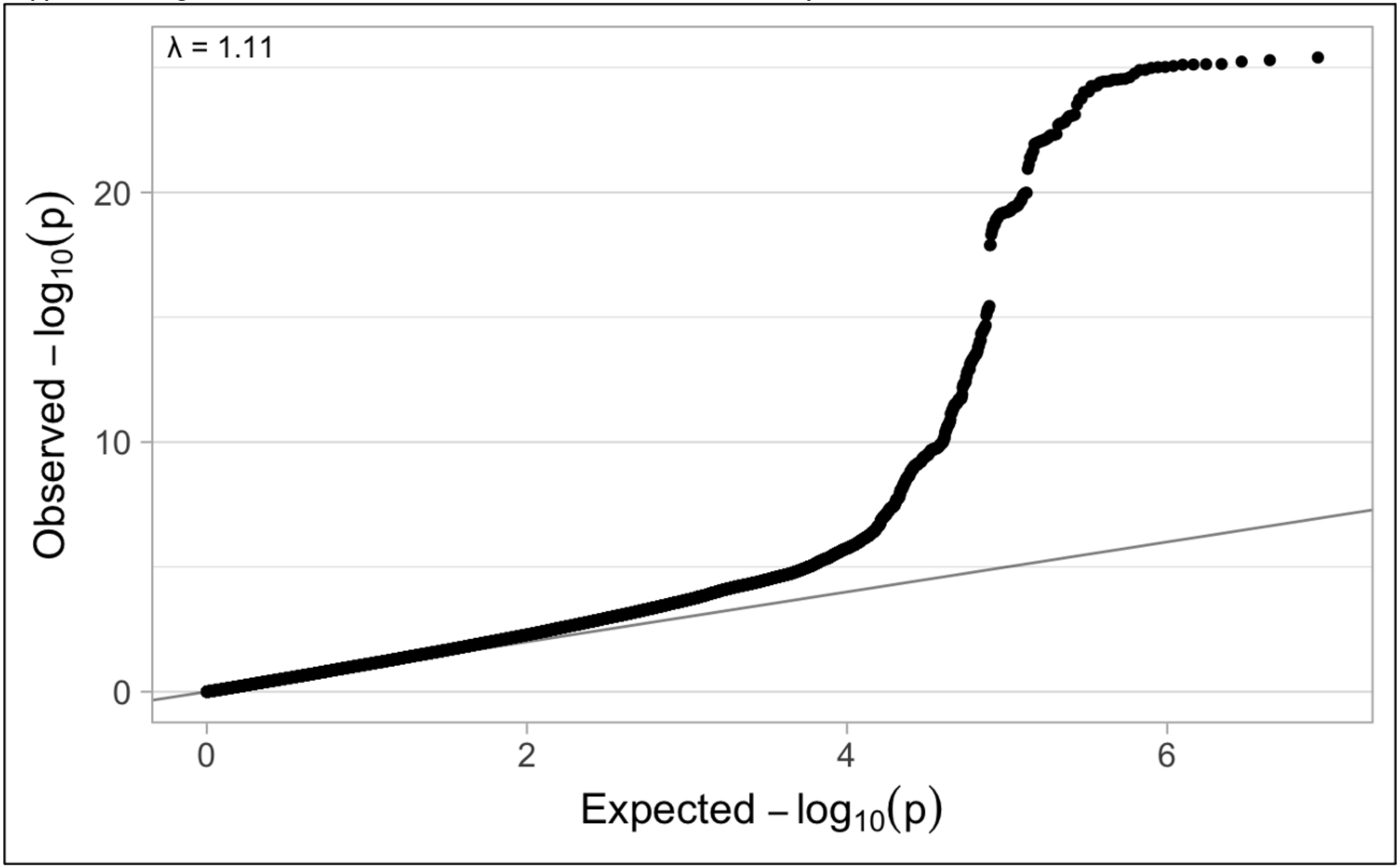


Quantile-quantile plot highlighting observed vs. expected p-values. LD Score Regression was performed to separate inflation of the test statistic from polygenicity or bias. LD Score intercept was 1.0059 (0.0062), consistent with inflation due to polygenicity.

**Supplemental Figure 3: N-GWAMA Regional Association Plots**

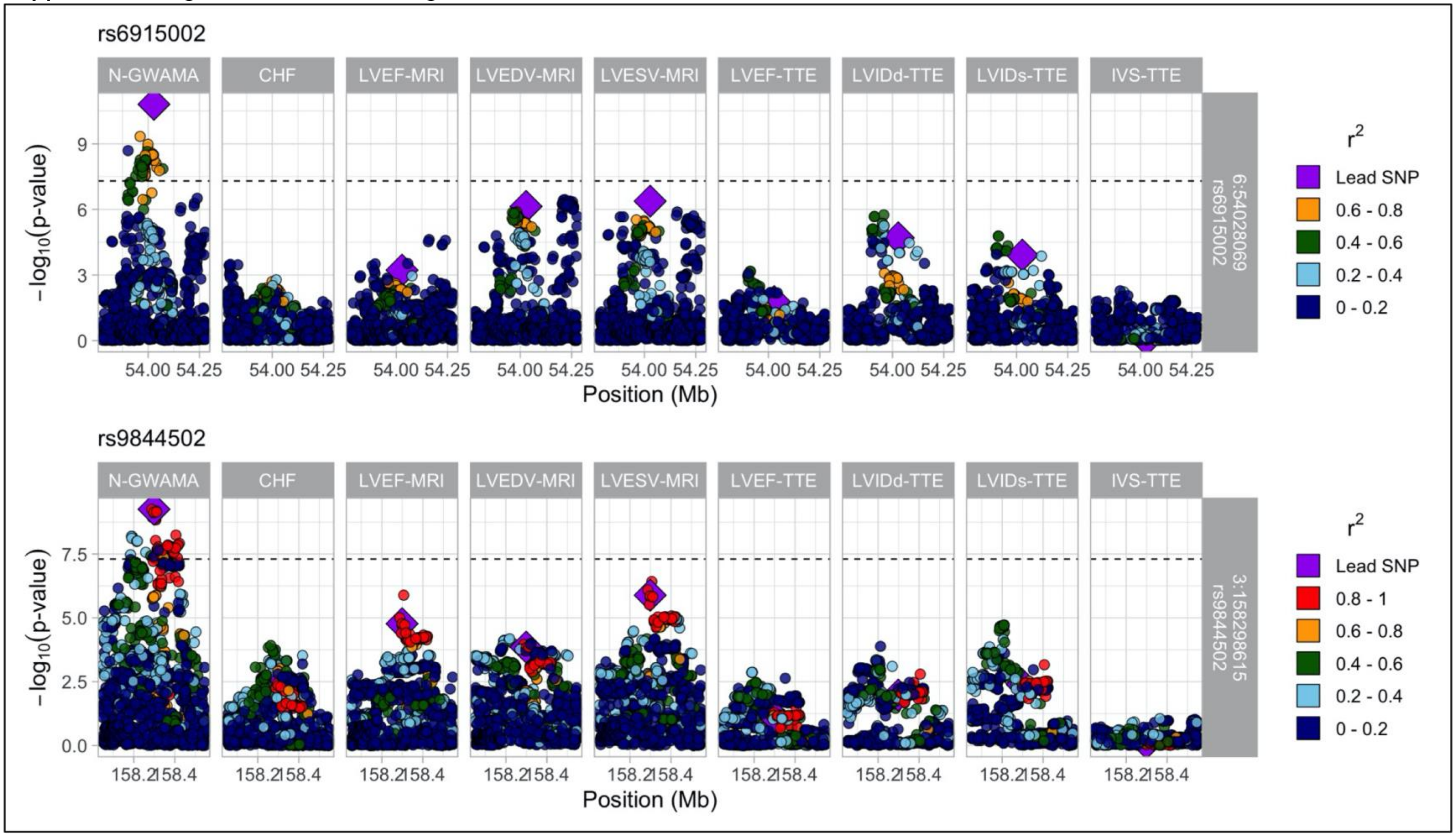

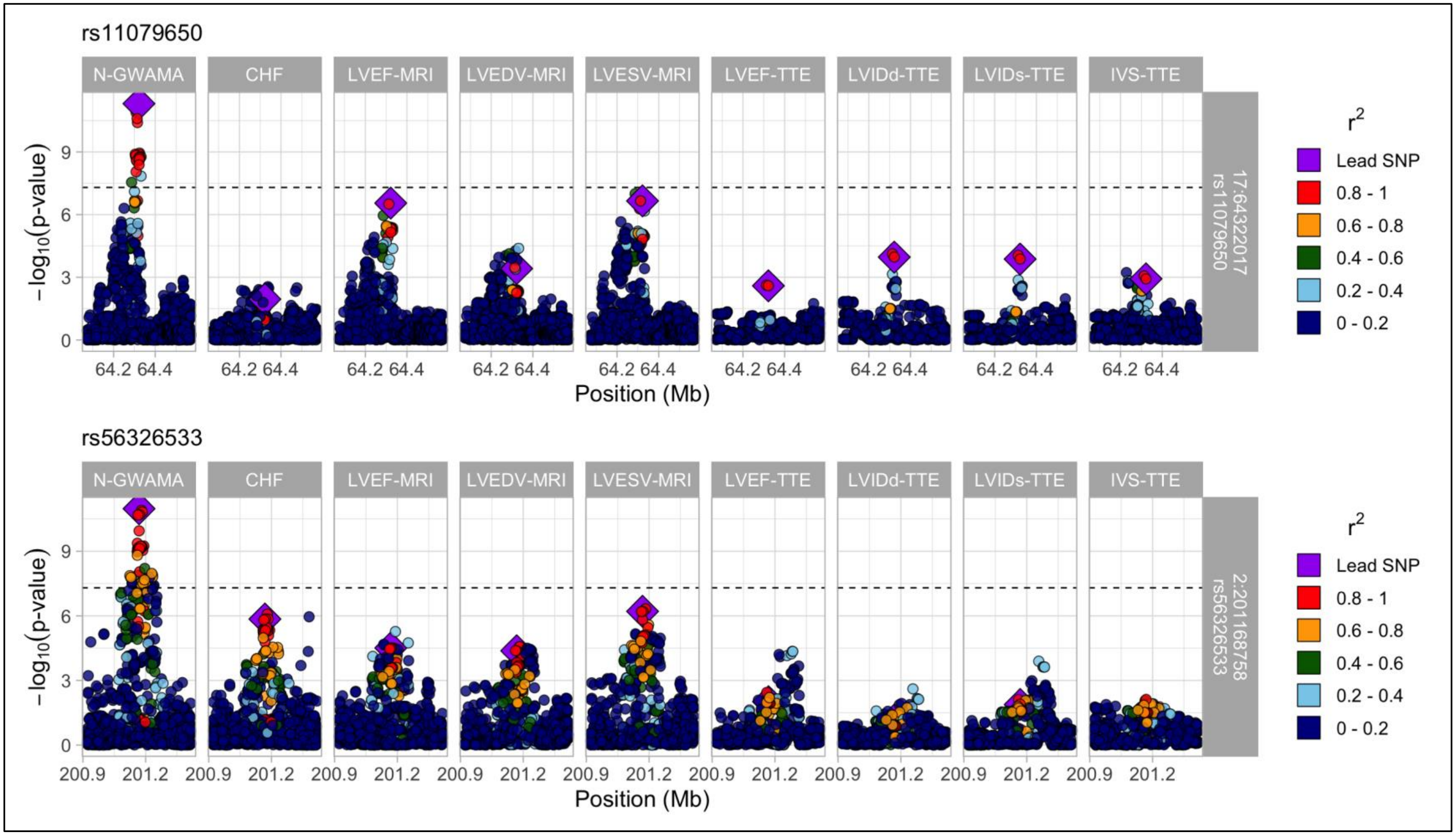
rs11079650
N-GWAMA
CHF
LVEF-MRI
LVEDV-MRI
LVESV-MRI
LVEF-TTE
LVIDd-TTE
LVIDs-TTE
IVS-TTE
17:64322017 rs11079650
−log10(p-value)
Position (Mb)
r2
Lead SNP
0.8 - 1
0.6 - 0.8
0.4 - 0.6
0.2 - 0.4
0 - 0.2
rs56326533
2:201168758 rs56326533

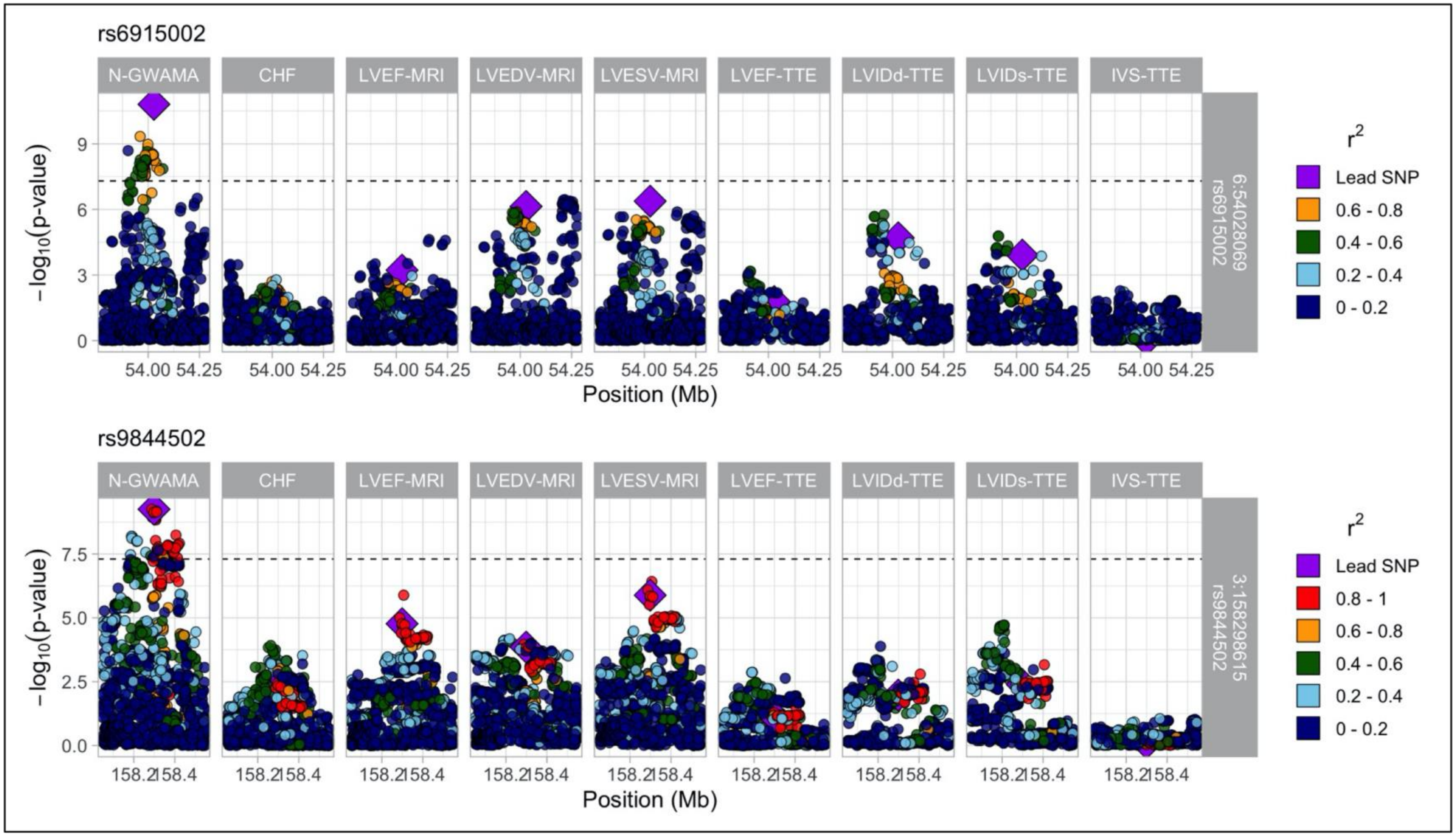

rs6915002
N-GWAMA
CHF
LVEF-MRI
LVEDV-MRI
LVESV-MRI
LVEF-TTE
LVIDd-TTE
LVIDs-TTE
IVS-TTE
−log10(p-value)
Position (Mb)
6:54028069
rs6915002
r2
Lead SNP
0.6 - 0.8
0.4 - 0.6
0.2 - 0.4
0 - 0.2
rs9844502
3:158298615
rs9844502
Lead SNP
0.8 - 1
0.6 - 0.8
0.4 - 0.6
0.2 - 0.4
0 - 0.2

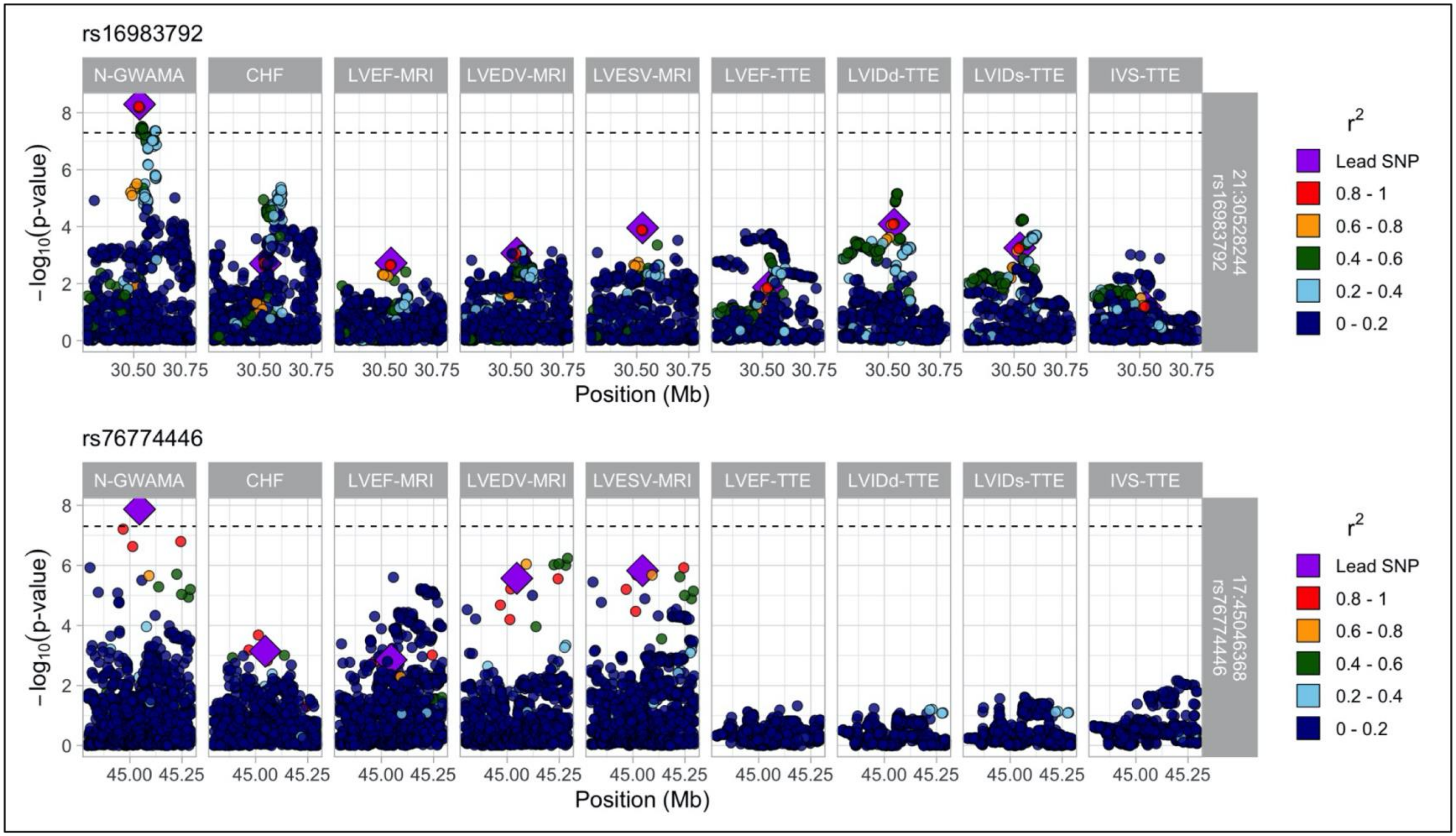
rs16983792
N-GWAMA
CHF
LVEF-MRI
LVEDV-MRI
LVESV-MRI
LVEF-TTE
LVIDd-TTE
LVIDs-TTE
IVS-TTE
21:30528244
rs16983792
−log10(p-value)
Position (Mb)
30.50 30.75
rs76774446
17:45046368
rs76774446
45.00 45.25
r2
Lead SNP
0.8 - 1
0.6 - 0.8
0.4 - 0.6
0.2 - 0.4
0 - 0.2

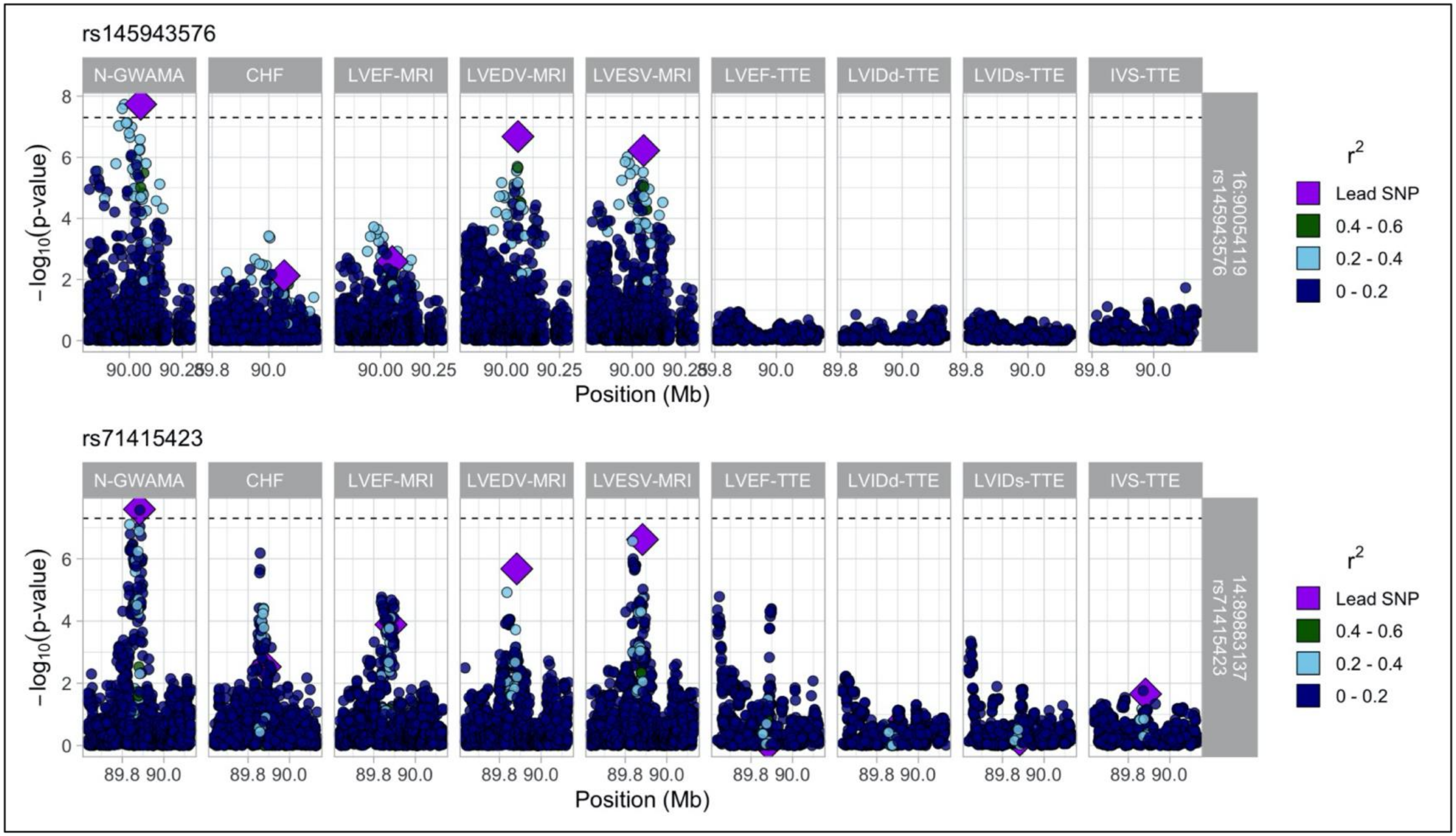

rs145943576
N-GWAMA
CHF
LVEF-MRI
LVEDV-MRI
LVESV-MRI
LVEF-TTE
LVIDd-TTE
LVIDs-TTE
IVS-TTE
16:90054119
rs145943576
−log10(p-value)
Position (Mb)
rs71415423
14:89883137
rs71415423
r2
Lead SNP
0.4 - 0.6
0.2 - 0.4
0 - 0.2

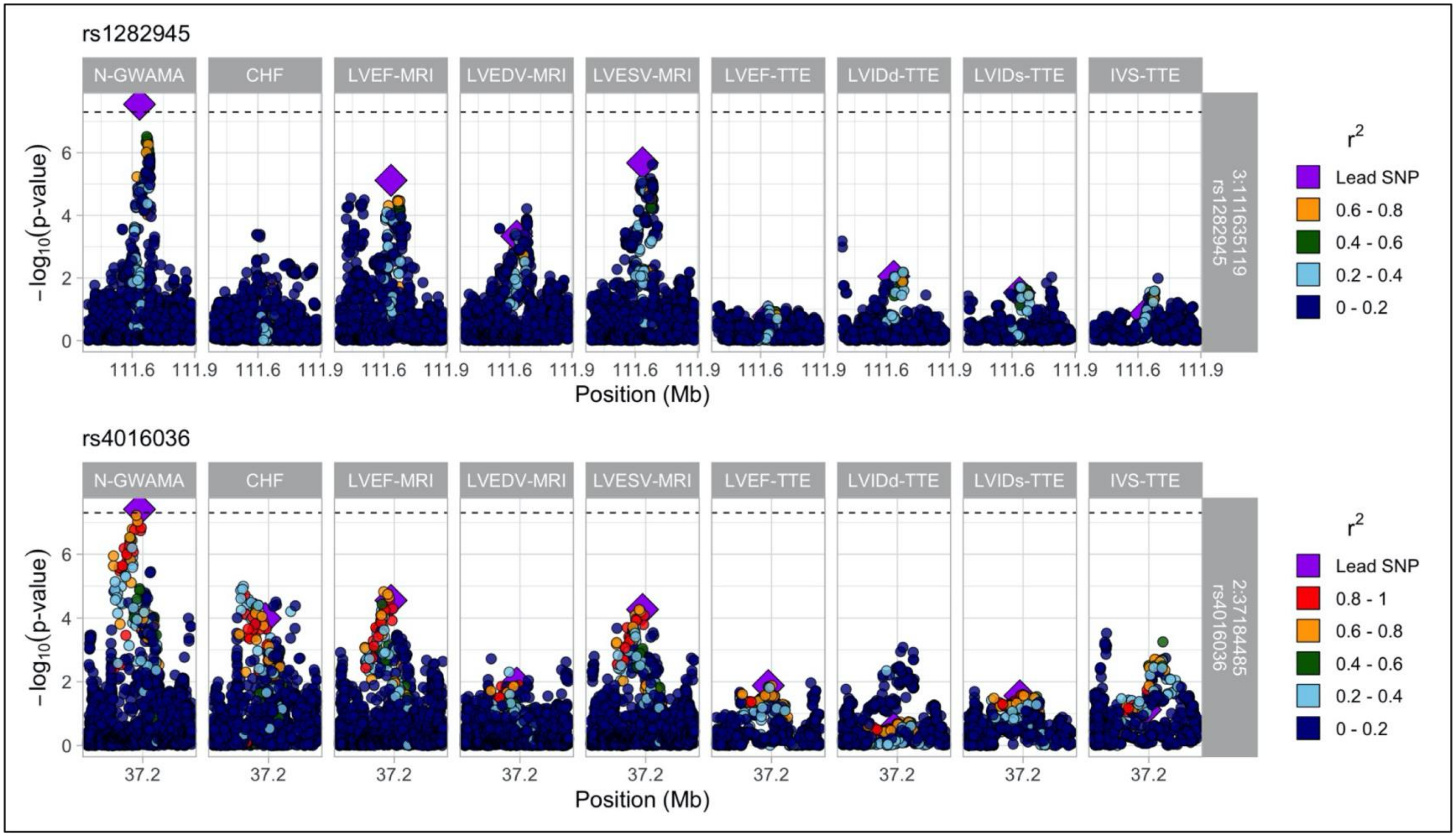
rs1282945
N-GWAMA
CHF
LVEF-MRI
LVEDV-MRI
LVESV-MRI
LVEF-TTE
LVIDd-TTE
LVIDs-TTE
IVS-TTE
3:111635119
rs1282945
−log10(p-value)
Position (Mb)
r2
Lead SNP
0.6 - 0.8
0.4 - 0.6
0.2 - 0.4
0 - 0.2
rs4016036
2:37184485
rs4016036
Lead SNP
0.8 - 1
0.6 - 0.8
0.4 - 0.6
0.2 - 0.4
0 - 0.2

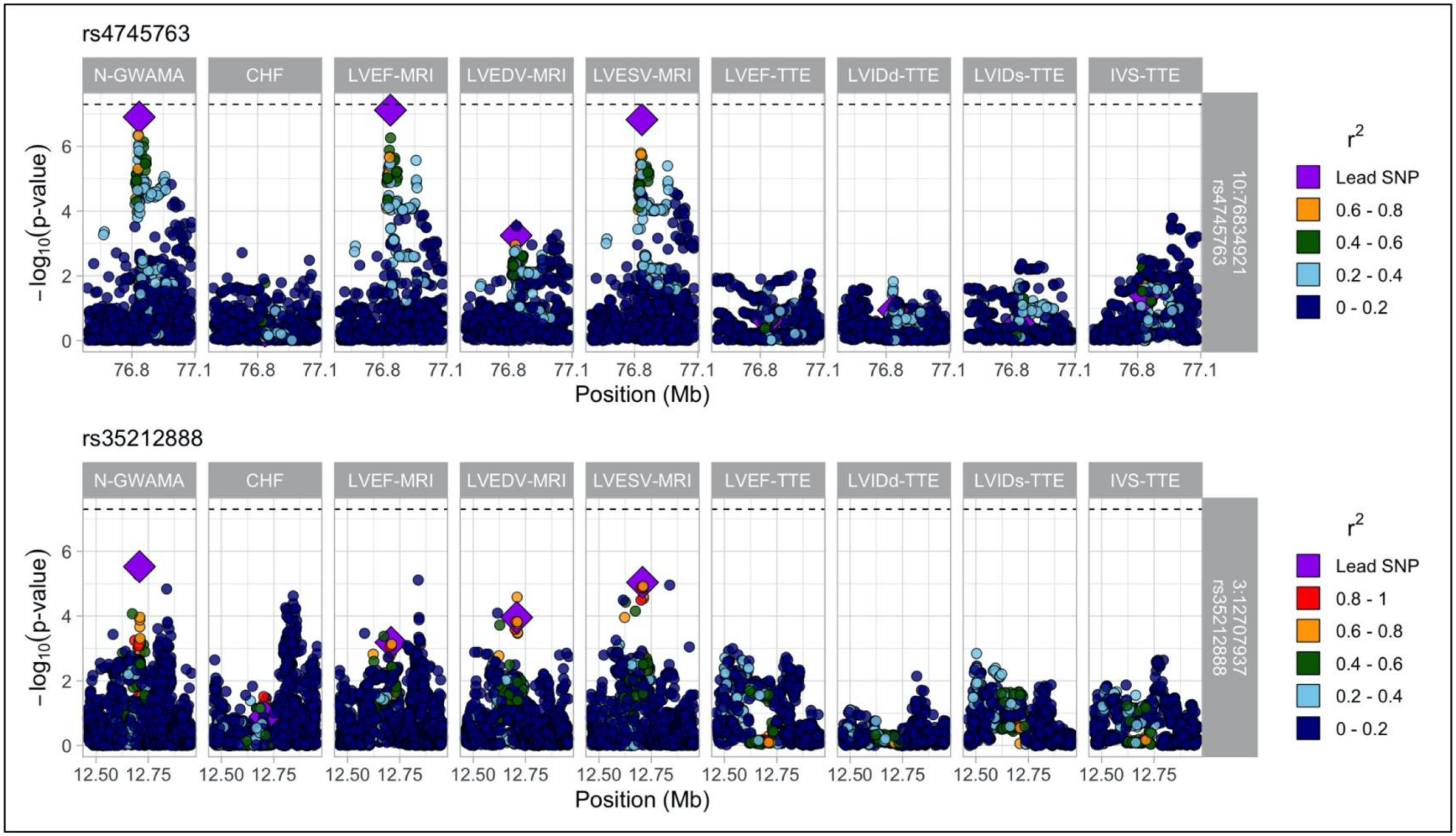
rs4745763
N-GWAMA
CHF
LVEF-MRI
LVEDV-MRI
LVESV-MRI
LVEF-TTE
LVIDd-TTE
LVIDs-TTE
IVS-TTE
10:76834921
rs4745763
−log10(p-value)
Position (Mb)
76.8
77.1
r2
Lead SNP
0.6 - 0.8
0.4 - 0.6
0.2 - 0.4
0 - 0.2
rs35212888
3:12707937
rs35212888
12.50
12.75
0.8 - 1

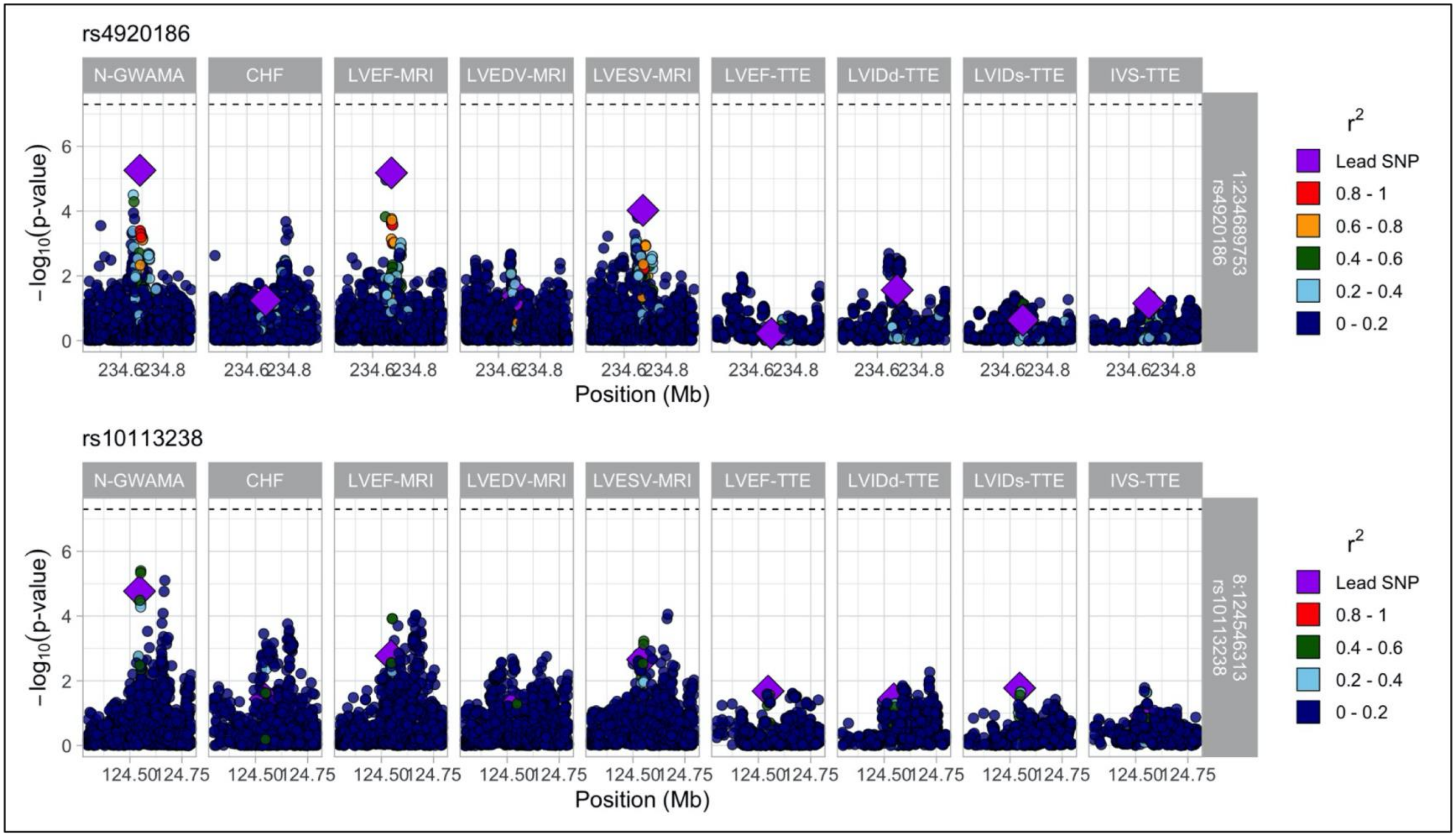
rs4920186
N-GWAMA
CHF
LVEF-MRI
LVEDV-MRI
LVESV-MRI
LVEF-TTE
LVIDd-TTE
LVIDs-TTE
IVS-TTE
1:234689753
rs4920186
−log10(p-value)
Position (Mb)
234.6
234.8
r2
Lead SNP
0.8 - 1
0.6 - 0.8
0.4 - 0.6
0.2 - 0.4
0 - 0.2
rs10113238
8:124546313
rs10113238
124.50
124.75

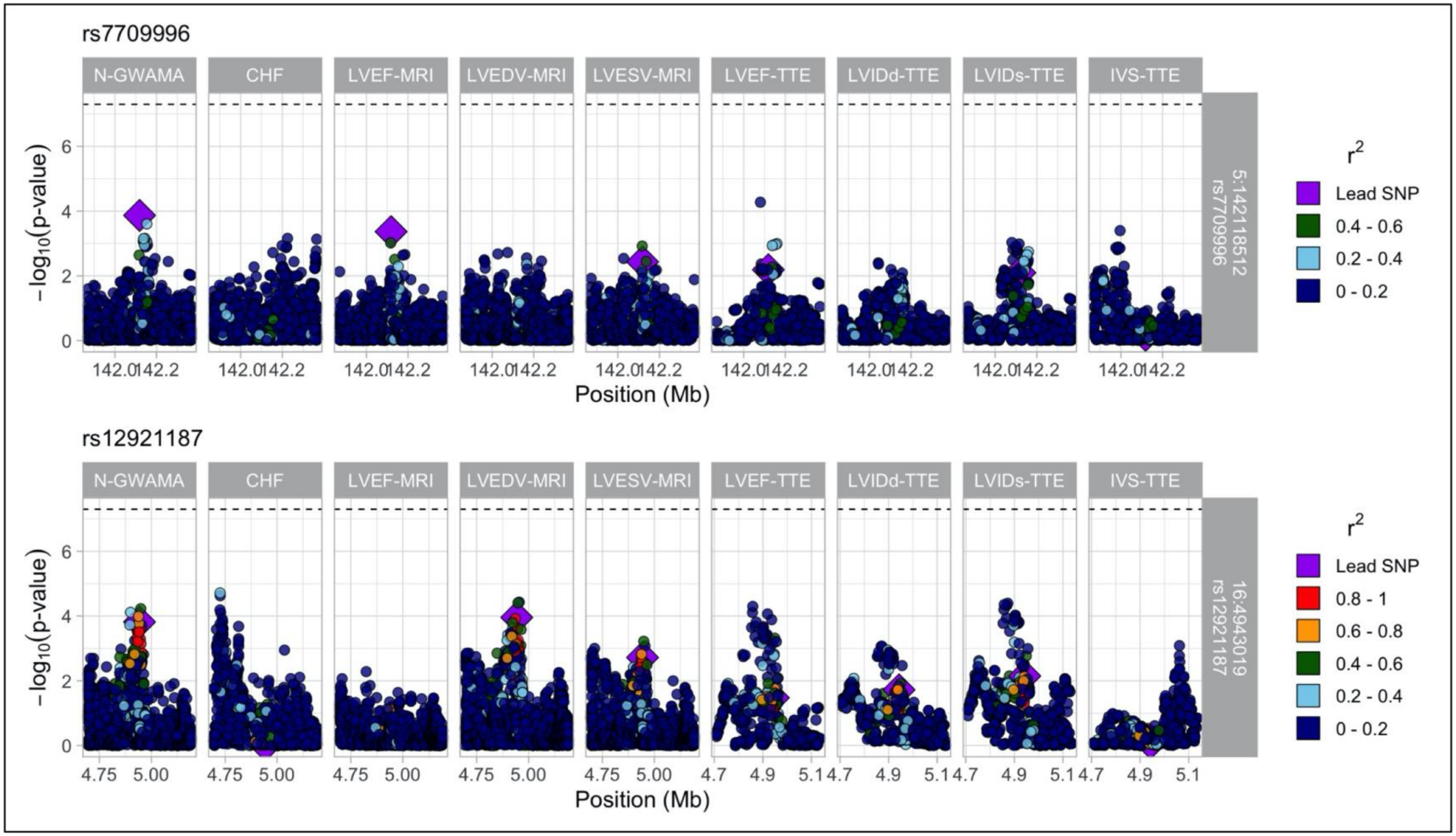


Regional association plots for novel conditionally-independent loci identified by N-GWAMA. Variants are plotted using unadjusted p-values from the initial N-GWAMA analysis (non-COJO). Colors of points represent linkage disequilibrium ($r^2$) with the lead SNP at each locus.

**Supplemental Figure 4: Overlap of Genome-wide Significant Variants Across GWAS Studies**

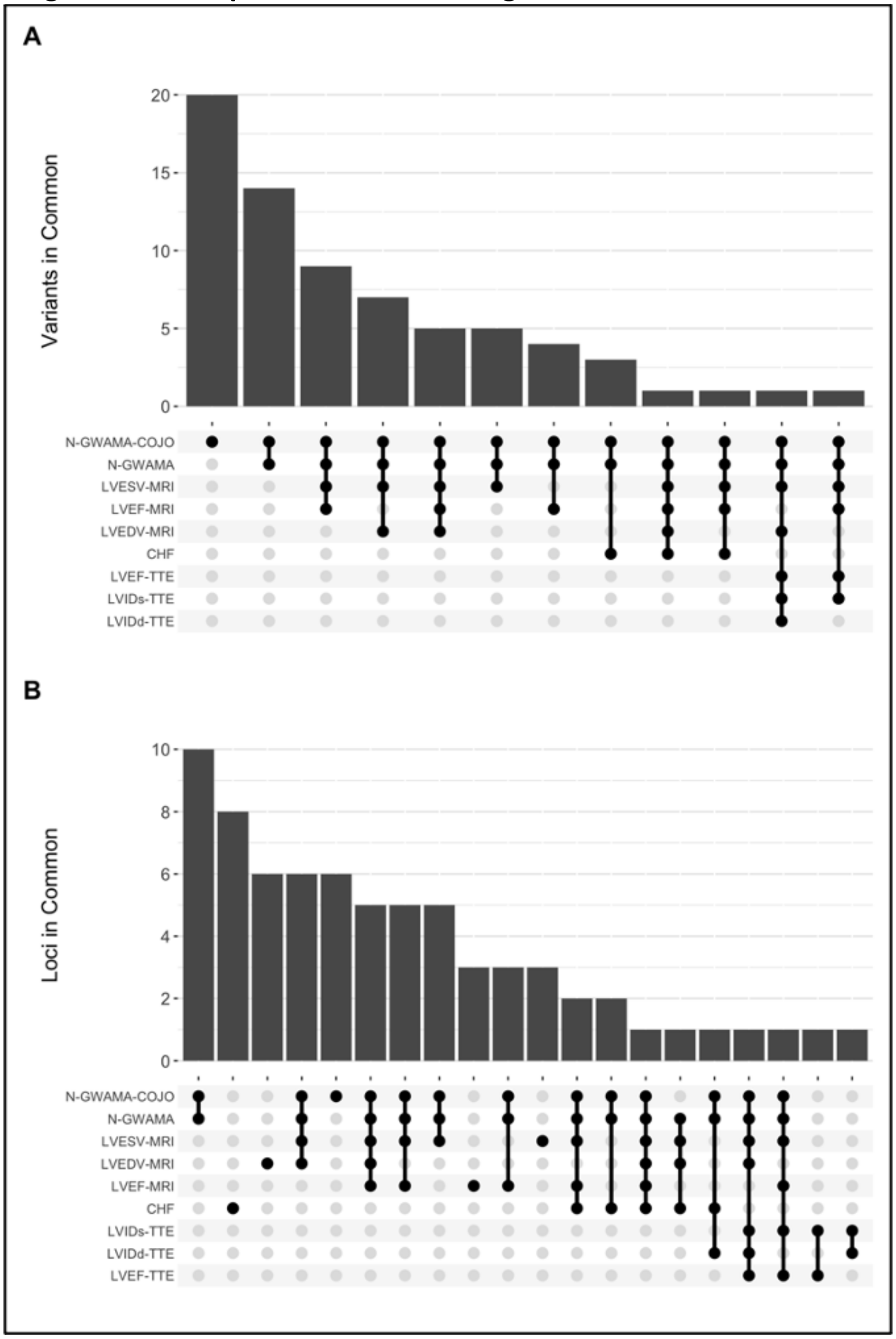


A) Number of variants and B) number of loci shared across genome-wide association studies of heart failure and cardiac imaging traits.

**Supplemental Figure 5: Enrichment of Cardiomyopathy Genes in GWAMA Loci**

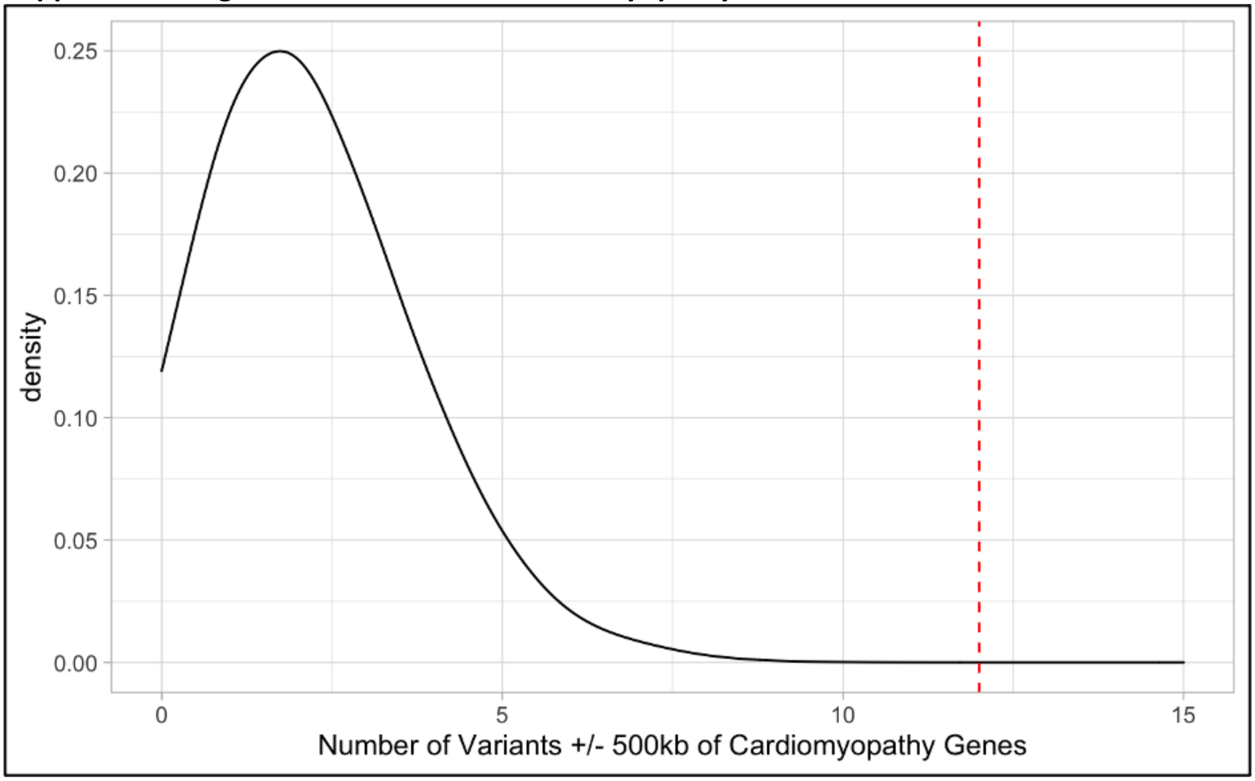


Enrichment of genetic variants located in/near genes implicated in Mendelian forms of cardiomyopathy (red dashed line) identified by N-GWAMA, in comparison to the distribution of matched SNPs across the genome identified using SNPsnap. We performed 10,000 permutations of matched SNPs to yield an empirical one-tailed p-value

**Supplemental Figure 6: Branch Chain Amino Acid Weighted Median Mendelian Randomization**

| Exposure | Outcome | n SNP | Method | Beta | 95% CI | P |
|---|---|---|---|---|---|---|
| Valine | LVESV-MRI | 10 | Weighted median | -0.00453 | [-0.078, 0.068] | 0.9 |
| Valine | LVEDV-MRI | 10 | Weighted median | 0.000424 | [-0.073, 0.073] | 1 |
| Leucine | LVESV-MRI | 5 | Weighted median | -0.122 | [-0.22, -0.028] | 0.01 |
| Leucine | LVEDV-MRI | 5 | Weighted median | -0.113 | [-0.21, -0.019] | 0.02 |
| Isoleucine | LVESV-MRI | 3 | Weighted median | -0.21 | [-0.33, -0.086] | $9 \times 10^{-4}$ |
| Isoleucine | LVEDV-MRI | 3 | Weighted median | -0.273 | [-0.4, -0.14] | $4 \times 10^{-5}$ |

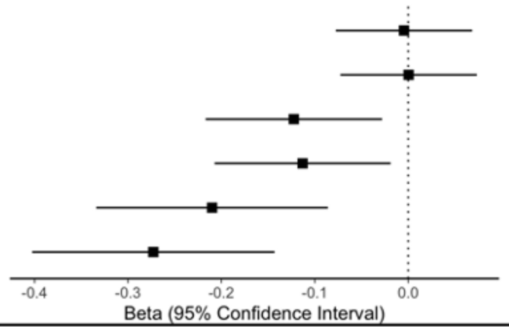


Mendelian randomization was performed to identify whether circulating branch chain amino acid levels were associated with cardiac MRI traits, given colocalization between BCKDHA and both $LVESV_{MRI}$ and $LVEDV_{MRI}$. Presented here are the results of weighted-median MR, which makes different assumptions about the presence of pleiotropy. This method remains robust when up to 50% of the weight of the genetic instrument is derived from invalid instruments.
